# FiMAP: A Fast Identity-by-Descent Mapping Test for Biobank-scale Cohorts

**DOI:** 10.1101/2021.06.30.21259773

**Authors:** Han Chen, Ardalan Naseri, Degui Zhi

## Abstract

Although genome-wide association studies (GWAS) have identified tens of thousands of genetic loci, the genetic architecture is still not fully understood for many complex traits. Most GWAS and sequencing association studies have focused on single nucleotide polymorphisms or copy number variations, including common and rare genetic variants. However, phased haplotype information is often ignored in GWAS or variant set tests for rare variants. Here we leverage the identity-by-descent (IBD) segments inferred from a random projection-based IBD detection algorithm in the mapping of genetic associations with complex traits, to develop a computationally efficient statistical test for IBD mapping in biobank-scale cohorts. We used sparse linear algebra and random matrix algorithms to speed up the computation, and a genome-wide IBD mapping scan of more than 400,000 samples finished within a few hours. Simulation studies showed that our new method had well-controlled type I error rates under the null hypothesis of no genetic association in large biobank-scale cohorts, and outperformed traditional GWAS approaches and variant set tests when the causal variants were untyped and rare, or in the presence of haplotype effects. We also applied our method to IBD mapping of six anthropometric traits using the UK Biobank data and identified a total of 3,442 associations, 2,224 (65%) of which remained significant after conditioning on independent association variants in the ± 3 cM flanking regions from GWAS.

## Introduction

Identity-by-descent (IBD) segments between two individuals are inherited from their common ancestor, without recombination.^1^ They have been widely used in forensic genetics,^2, 3^ as well as population genetics to detect evidence of natural selection, and to estimate the demographic history such as bottlenecks and admixture.^4-6^ In a genome-wide association study (GWAS) with cryptically related samples, the average proportion of the genome shared IBD can be used to infer the degree of relatedness,^1, 7^ and to construct an empirical kinship matrix to account for cryptic relatedness in GWAS.^8-11^

IBD mapping is the study of association between the sharing of IBD segments and phenotypic similarities. Early IBD mapping studies were mostly in linkage analysis for family studies.^12-15^ IBD mapping methods for other study designs have also been developed, such as those searching for chromosome segments shared by distantly related patients,^16^ testing for association with haplotype clusters,^17^ and comparing pairwise IBD rates in case-case and case-control pairs.^18^ Indeed, IBD mapping has successfully identified genomic regions associated with Parkinson’s disease,^19^ serum triglycerides,^20^ multiple sclerosis,^21^ and amyotrophic lateral sclerosis.^22^

Although IBD mapping is not yet a widely-applied method, it holds promise to uncover untyped rare variant and haplotypic effects, that are still escaping the search of the “missing heritability”. While most common variants are tagged by genotypes from arrays and can be well-imputed, rare variants are still not well covered. IBD mapping can indirectly test the effects of rare variants co-segregated with the IBD segments and recover the association signal. Further, the combination of multiple variants on an IBD segment, each individually weakly associated with the phenotype and thus difficult to be identified by single variant association tests, can be captured by IBD mapping. As the phase information is often ignored in traditional GWAS approaches and variant set tests for rare genetic variants,^23-29^ IBD mapping leverages such information and is less susceptible to genotyping or sequencing errors. It can better identify association signals from rare variants and haplotype effects,^18^ and therefore offers a unique angle to investigate genetic associations.

While promising, IBD mapping were not popular due to several technical challenges. First, IBD mapping requires accurate phasing of haplotypes. This difficulty is mostly addressed by current phasing software.^30-32^ Second, the power of IBD mapping methods is linked with the number of IBD segments in the sample. While the density of IBD segments in families and inbred populations are high, their density in outbred populations can be much lower. Nonetheless, with the availabilities of large biobanks, the sample sizes are large enough to harbor a large number of IBD segments. For example, our recent studies found that each haplotype of a UK Biobank participant is covered by about 10 5cM IBD segments shared with other UK Biobank participants.^33^ Third, traditional IBD segment calling methods are not efficient and cannot scale up to modern biobank-scale cohorts with hundreds of thousands of samples. Fortunately, this challenge is largely resolved by a new generation of efficient IBD segment detection methods, either based on the positional Burrows Wheeler transform (PBWT) algorithm,^34^ such as RaPID,^35^ hap-IBD,^36^ and TPBWT,^37^ or based on advanced string hashing such as FastSMC^38^ and iLASH.^39^ These methods typically achieve *O*(*N*) complexity, where *N* is the sample size, and have made IBD segment calling computationally feasible in biobank-scale cohorts with hundreds of thousands to millions of individuals. However, even with the advancements of abovementioned technologies, existing IBD mapping methods are still not scalable to large sample sizes due to lack of efficient statistical tests for IBD mapping.

To address the main computational challenge for biobank-scale cohorts that any IBD mapping methods with *O*(*N*^2^) computational time complexity or higher would quickly become infeasible, we have developed FiMAP, a fast IBD mapping test for biobank-scale cohorts. FiMAP leverages IBD segments identified by RaPID^35^ and constructs a genotype similarity tensor on the whole genome (**Figure 1**). The genotype similarity tensor is a collection of *N* × *N* pairwise local IBD matrices that change across the whole genome. Each local IBD matrix is a sparse matrix indicating the proportion of IBD sharing between any two individuals in the given genomic region, with most elements being 0. The average of local IBD matrices across the whole genome is the global IBD matrix representing the average proportion of IBD sharing, which can be used as a kinship coefficient matrix to account for genetic relatedness in the study samples. FiMAP leverages a random matrix-based algorithm to speed up the computation, so that a genome-wide IBD mapping analysis for hundreds of thousands of study samples can be finished within a few hours. We also demonstrate the application of FiMAP to the IBD mapping analysis of six anthropometric traits in the UK Biobank.

**Figure 1.**
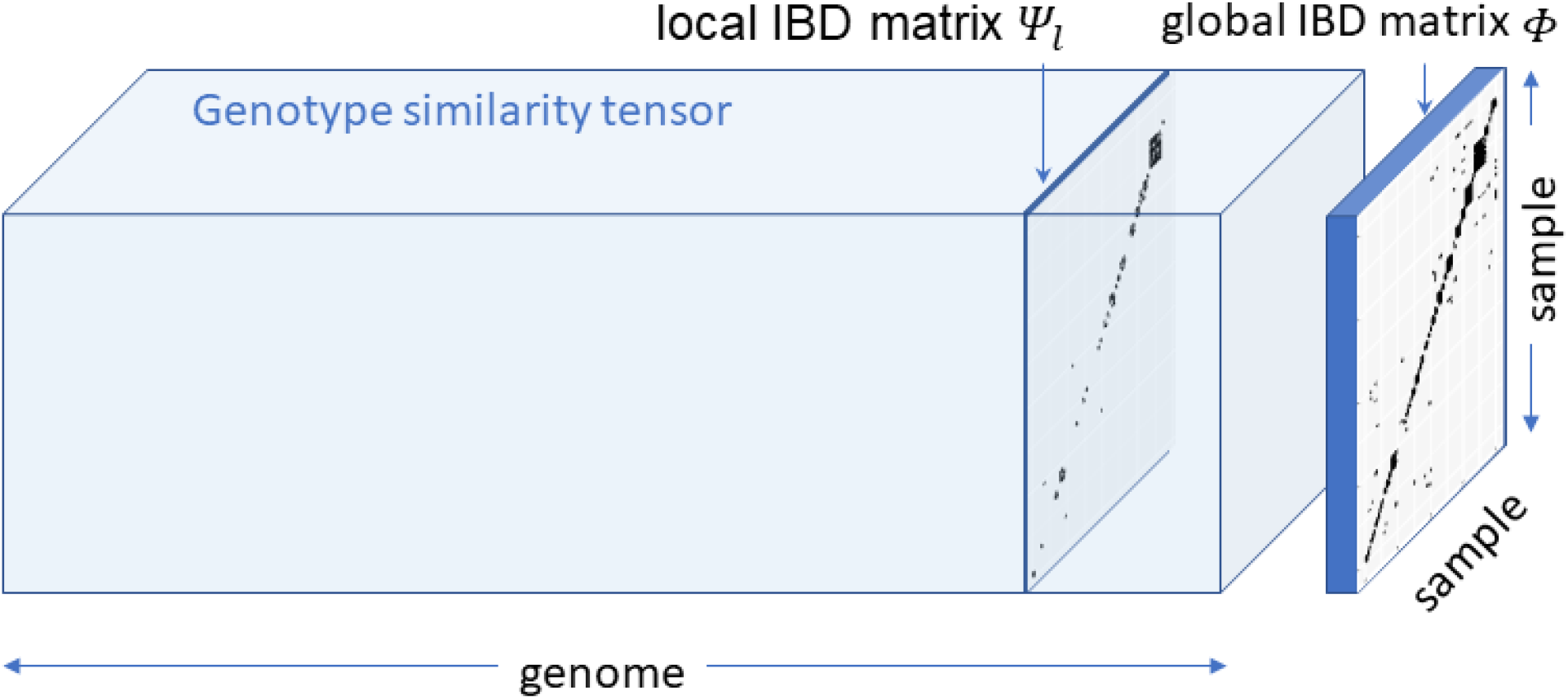
Local and global IBD matrices in FiMAP. The global IBD matrix *Φ* is the average of the local IBD matrices *Ψ*_*l*_ along the genome dimension of the genotype similarity tensor.

## Methods

### Variance component models

Variance component models have long been used in linkage analysis to identify genetic loci linked with quantitative traits.^12-15^ Consider a linear mixed model *Y*_*i*_ = *X*_*i*_*β* + *b*_*i*_ + *δ*_*il*_ + *ε*_*i*_ for individual *i*, where *Y*_*i*_ is the phenotype, *X*_*i*_ are *c* fixed-effect covariates with effect sizes *β, b*_*i*_ is the polygenic random effect accounting for global relatedness, *δ*_*il*_ is the random effect of local relatedness at genetic locus *l*, we stack *N* individuals to get length *N* vectors *Y, b, δ*_*l*_, *ε*, and an *N* × *c* matrix *X*, from *Y*_*i*_, *b*_*i*_, *δ*_*il*_, *ε*_*i*_, *X*_*i*_, respectively. The vector 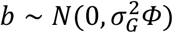 are the random effects for the *N* × *N* global IBD (or kinship) matrix *Φ* denoting the average proportion of IBD sharing between any two individuals, 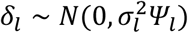 are the random effects for the *N* × *N* local IBD matrix *Ψ*_*l*_ at this genetic locus *l*, and *ε* are independent and identically distributed errors that follow a normal distribution with mean 0 and variance 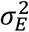. Under the null hypothesis of no association with the quantitative trait at genetic locus *l*, the random effects *δ*_*l*_ are equal to 0 and this is equivalent to testing variance component hypotheses 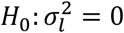 vs 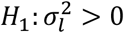.

### The asymptotic test

Let 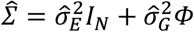, where both 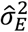 and 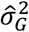 are estimated from the null model (with *δ*_*l*_ = 0) in the matrix-vector form *Y* = *Xβ* + *b* + *ε*. Let 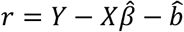 be the length *N* residual vector, where both 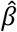 and 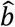 are estimated from this null model, the classical score-type variance component test 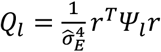 asymptotically follows a chi-square mixture distribution Σ_*j*_ ζ_*j*_ χ_1,*j*_ ^2,27, 40, 41^ where χ_1,*j*_ ^2^ are independent chi-square distributions with 1 degree of freedom, and ζ_*j*_’s are the eigenvalues of 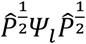, where 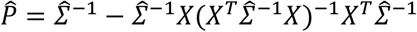 is the *N* × *N* projection matrix.^42^

### The finite-sample adjustment

Although under regularity conditions, 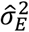 and 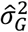 are consistent estimators for variance component parameters 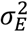 and 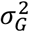 under the null hypothesis 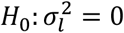, the classical score-type variance component test above treats them as fixed numbers and ignores the variability in their estimation, which could result in not well-calibrated p values in finite samples. This is a known issue for score-type variance component tests in microbiome association studies with small sample sizes.^43-45^ Despite large sample sizes in biobank-scale cohorts, the local IBD matrix *Ψ*_*l*_ for genetic locus *l* is often sparse, which could invalidate asymptotic inference on the quadratic form 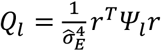. We note that 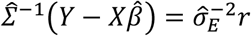, and 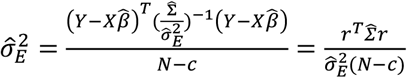, where 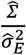 is free of 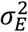. To account for the variability in estimating 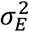, we can rewrite 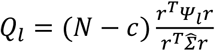, and compute the finite-sample p-value as *P*(Σ_*j*_ *ξ*_*j*_χ_1,*j*_^2^ > 0), where *ξ*_*j*_’s are the eigenvalues of 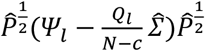.

### FiMAP

As both 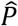 and *Ψ*_*l*_ are *N* × *N* matrices (and 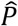is not sparse), conducting the classical score-type variance component test, without or with the finite-sample adjustment, requires *O*(*N*^2^) memory footprint and *O*(*N*^3^) computational time complexity, which becomes infeasible for hundreds of thousands to millions of individuals. To solve this computational challenge, we use an *N* × *B* random matrix *R* to compute approximated top *B* eigenvalues of 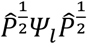 from 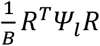.

Specifically, we start with a Cholesky decomposition of the sparse global IBD matrix *Φ* = *LL*^*T*^, then simulate length *N* random vectors *r*_1_ and *r*_2_ from a standard normal distribution to compute 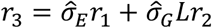 and 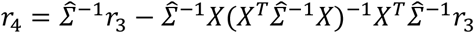. Therefore, we have 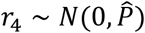 and repeat the process *B* times to get an *N* × *B* random matrix *R*. Assuming both the global IBD matrix *Φ* and the local IBD matrices *Ψ*_*l*_ are block-diagonal (with a bounded largest block size) and sparse, then both *L* and 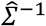 are also block-diagonal, and *r*_3_ and *r*_4_ do not require any matrix-vector multiplications involving full dense *N* × *N* matrices. If both the largest block size and *B* are *O*(1), then the overall computational complexity of FiMAP is *O*(*N*). Moreover, for the finite-sample adjustment, we can pre-compute 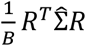 only once in a genome-wide scan, and approximate top *B* eigenvalues 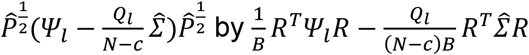.

In reality, we observe the start and end positions of each IBD segment between two individuals (e.g., from RaPID^35^). Therefore, for a genetic locus *l*, instead of a local IBD matrix *Ψ*_*l*_ with 2 identical chromosomes for each individual with themselves, we construct 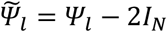, with each element denoting the proportion of IBD segments between two individuals in the given region. For biobank-scale cohorts, 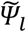 is highly sparse with most elements 0. Assuming a total of *L* equally spaced genomic regions across the whole genome, the global IBD matrix 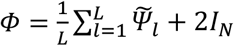 is the average of the local IBD matrices *Ψ* ‘s. In our implementation, we pre-compute an offset 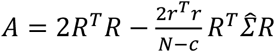 only once in a genome-wide scan. For each genetic locus *l*, we then compute the finite-sample p-value as 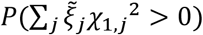, where 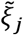’s are the eigenvalues of 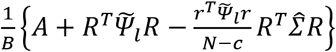. Note that 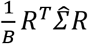 is also pre-computed, for each genetic locus *l* with observed IBD proportion matrix 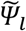, we only need to update the *B* × *B* matrix 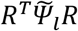 and the scalar 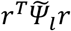.

### Simulation studies

We used RaPID and hap-IBD IBD segment calls from the UK Biobank array-typed genotype data to simulate phenotype data, and evaluated type I error rates and power of FiMAP in identifying genetic associations. Specifically, we used RaPID IBD segment calls on all 22 autosomes to construct the global IBD matrix *Φ* for 487,252 individuals. For individuals with no inbreeding, the diagonal elements of *Φ* is equal to 2, which is 4 times the theoretical kinship coefficient, indicating the total length of IBD segments shared by each individual with themselves is twice the total length of all autosomes. The off-diagonal elements of *Φ* are the total length of IBD segments shared by each pair of individuals, divided by the total length of all autosomes. To ensure the sparsity of the global IBD matrix *Φ*, we set off-diagonal elements less than 0.088 to 0, which is equivalent to including fourth-degree relatives or closer in the kinship matrix (kinship coefficient ≥ 0.022). We then chunked 22 autosomes into 1 cM windows and assembled 3,403 local IBD matrices by computing the proportion of IBD sharing between each pair of individuals in each 1 cM window (the last window on each chromosome is shorter than 1 cM). We set the number of random vectors used in FiMAP *B* = 100 in all simulation studies.

In type I error simulations, we randomly selected 400,000 individuals and subset the global IBD matrix *Φ* to get a submatrix *Φ*_0_. We then simulated age from a normal distribution with mean 50 and standard deviation 5, and sex from a Bernoulli distribution with probability 0.5. A continuous phenotype *Y*_*i*_ for individual *i* was simulated as *Y*_*i*_ = 0.05*age*_*i*_ + 0.5*sex*_*i*_ + *b*_*i*_ + *ε*_*i*_, where the vector form *b* followed a multivariate normal distribution with mean 0 and covariance matrix *Φ*_0_, and *ε*_*i*_ followed a standard normal distribution. Then we tested for the association with 3,403 local IBD matrices after adjusting for age and sex. We simulated 50 phenotype replicates and obtained a total of 170,150 p values under the null hypothesis of no local IBD random effects.

We also conducted power simulations to benchmark finite-sample FiMAP results with standard GWAS results as well as variant set tests, including the burden test,^26^ SKAT,^27^ SKAT-O,^28^ and SMMAT^46^ on the same 1 cM window. Using a random sample of N = 400,000 individuals from the UK Biobank, we simulated untyped ultra-rare causal variants that were not included in any of the tests for a fair comparison. Specifically, in each simulation replicate, we randomly selected a 1 cM window with at least 2 ultra-rare variants with MAF < 0.05%, we then assumed that all *J* ultra-rare variants with MAF < 0.05% in this window were causal variants and simulated a continuous phenotype *Y*_*i*_ for individual *i* as 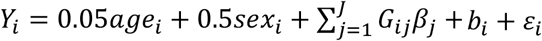, where *age*_*i*_, *sex*_*i*_, *b*_*i*_ and *ε*_*i*_ were simulated using the same parameter settings as in the type I error simulations, and the causal variant effect 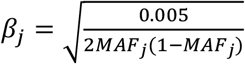. Finite-sample FiMAP with local IBD matrices constructed from RaPID and hap-IBD IBD segments called with length ≥ 3 cM, 5 cM, 10 cM were conducted. Using the directly genotyped data, we excluded variants with missing rate > 5% or MAF < 1% to mask the ultra-rare causal variants, and conducted a standard single-variant test commonly used in GWAS. We selected the minimum single-variant test p value in the same 1 cM window and applied a Bonferroni correction to account for multiple testing. We also conducted four variant set tests on the same genotyped data as used in the single-variant test, in the same 1 cM window: the burden test, SKAT, SKAT-O and SMMAT, all using the same Wu weights.^27^ Empirical power was estimated as the proportion of p values (for the single-variant test, Bonferroni-corrected p values) less than 0.05 over 1,000 simulation replicates.

In addition, we simulated haplotype effects and compared FiMAP with IBD segments called with length ≥ 3 cM, 5 cM, 10 cM with the single-variant test as well as the aforementioned four variant set tests. We computed all haplotypes on autosomes, with length ≥ 0.5 cM shared by 500 or more individuals from the UK Biobank using a PBWT-block algorithm.^47^ In each simulation replicate, we used a random sample of N = 400,000 individuals and randomly sampled a causal haplotype. We simulated a continuous phenotype *Y*_*i*_ for individual *i* as *Y*_*i*_ = 0.05*age*_*i*_ + 0.5*sex*_*i*_ + *H*_*i*_*γ* + *b*_*i*_ + *ε*_*i*_, where *age*_*i*_, *sex*_*i*_, *b*_*i*_ and *ε*_*i*_ were simulated using the same parameter settings as in the type I error simulations, and *H*_*i*_ was the number of causal haplotypes carried by individual *i*, with possible values 0, 1, 2. The causal haplotype effect was assigned as 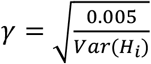. We then tested the 1 cM window with the largest overlap with the causal haplotype using FiMAP, single-variant test and variant set tests.

### UK Biobank anthropometric traits

We applied FiMAP to 6 anthropometric traits: waist circumference (N = 407,872), hip circumference (N = 407,827), standing height (N = 407,681), sitting height (N = 407,323), body mass index (N = 407,255), and body weight (N = 407,400), from the UK Biobank white British study participants with genetic ethnic group in Caucasians, after excluding individuals with inconsistent gender and biological sex. Each trait was adjusted for age, age^2^, sex, their interactions, and top 10 ancestral principal components (PCs), and the residuals were rank normalized and analyzed using a linear mixed model with the same aforementioned fixed-effects covariates^48^ and the global IBD matrix to model the covariance structure of the random intercept. We used RaPID and hap-IBD IBD segment calls with length ≥ 3 cM, 5 cM, 10 cM, and defined genomic regions *l* using 1 cM windows as used in our simulation studies, with a total of 3,403 test regions on 22 autosomes. We set the number of random vectors used in FiMAP *B* = 100 in all UK Biobank data analyses. For each trait, genome-wide significance was defined as finite-sample FiMAP p value < 0.05/3,403 = 1.47 × 10^−5^, the Bonferroni-corrected threshold for 3,403 tests.

We also analyzed the same anthropometric traits in a GWAS setting using GMMAT^42^ on imputed genetic variants with MAF ≥ 0.01%, imputation quality score ≥ 0.3, and missing rate < 5%. For each trait, genome-wide significance was defined as single-variant test p value < 5 × 10^−8^. In the conditional analysis, for each 1 cM window with a significant finite-sample FiMAP p value < 1.47 × 10^−5^, we performed stepwise model selection to identify a set of imputed genetic variants with GWAS p values that reached genome-wide significance in the ± 3 cM flanking regions (a total length of 7 cM) after conditioning on each other, and then adjusted for this set of imputed genetic variants in the conditional FiMAP analysis.

## Results

### Type I error rates

We first counted the numbers of non-zero off-diagonal elements in each of the 3,403 local IBD matrices for 1 cM windows, with sample size N = 487,252. The local IBD matrices were constructed from RaPID and hap-IBD IBD segments called with length ≥ 3 cM, 5 cM, 10 cM. **Figure 2** shows that the number of non-zero off-diagonal elements increased as the IBD length cutoff decreased from 10 cM, 5 cM to 3 cM, for IBD segments called from both RaPID and hap-IBD. For example, when the RaPID IBD length cutoff was 3 cM, the median number of non-zero elements per individual in local IBD matrices from the full sample was 60.4, with a range of 8.2 to 468.0. In contrast, in a random subset of N = 1,000 individuals, the median number of non-zero elements per individual was 0.124, with a range of 0.021 to 1.326. For hap-IBD, the maximum number of non-zero elements per individual in local IBD matrices with the IBD length cutoff of 3 cM was 277× the sample size, suggesting that the number of non-zero elements in any local IBD matrix at a given length cutoff (≥ 3 cM, 5 cM, or 10 cM) in the UK Biobank data was about a constant factor of *N*, regardless of the IBD caller, thus the computational efficiency of FiMAP was guaranteed. On a computing server with dual Intel^®^ Xeon^®^ E5-2687W v4 CPU (3.00 GHz, 24 cores in total), analysis of each simulation replicate with N = 400,000 took about 131 minutes for RaPID IBD length cutoff 3 cM, 36 minutes for 5 cM, and 9.4 minutes for 10 cM, using 40 threads in parallel.

**Figure 2.**
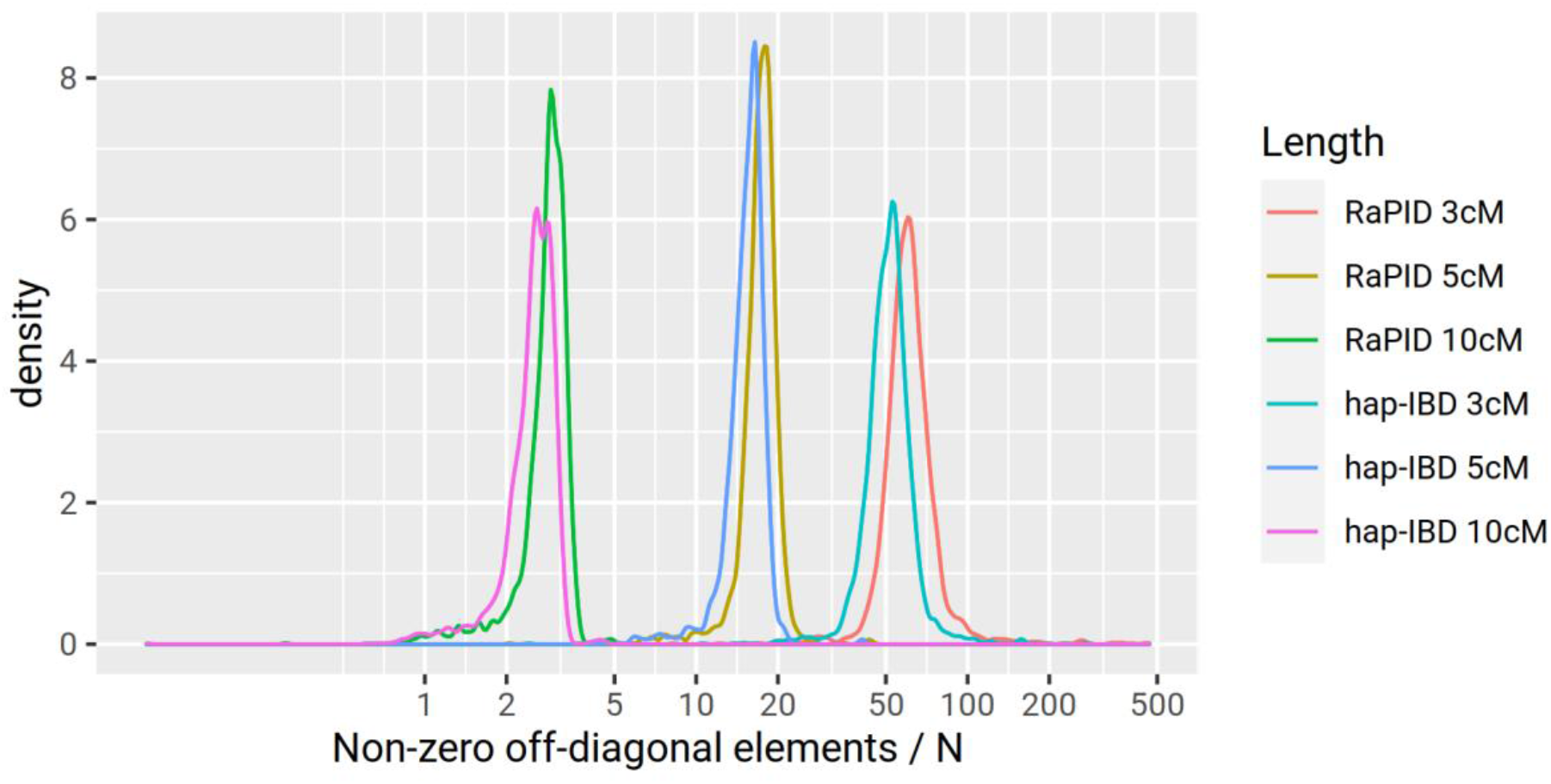
Distribution of average numbers of non-zero IBD sharing per individual in local IBD matrices. For each color, a total of 3,403 local IBD matrices of sample size N = 487,252 for 1 cM windows were plotted. The local IBD matrices were constructed from RaPID and hap-IBD IBD segments called with length ≥ 3 cM, 5 cM, 10 cM.

Asymptotic and finite-sample FiMAP p values under the null hypothesis of no local IBD random effects were shown in **Figure 3**. Asymptotic FiMAP p values were extremely conservative in the tail, while FiMAP p values with the finite-sample adjustment were well-calibrated, for IBD segments called from both RaPID and hap-IBD. The median finite-sample FiMAP p value showed genomic inflation factors very close to 1 for IBD segments called with length ≥ 3 cM, 5 cM, 10 cM, suggesting that FiMAP with the finite-sample adjustment is a fast and valid IBD mapping test in large samples, regardless of the IBD caller.

**Figure 3.**
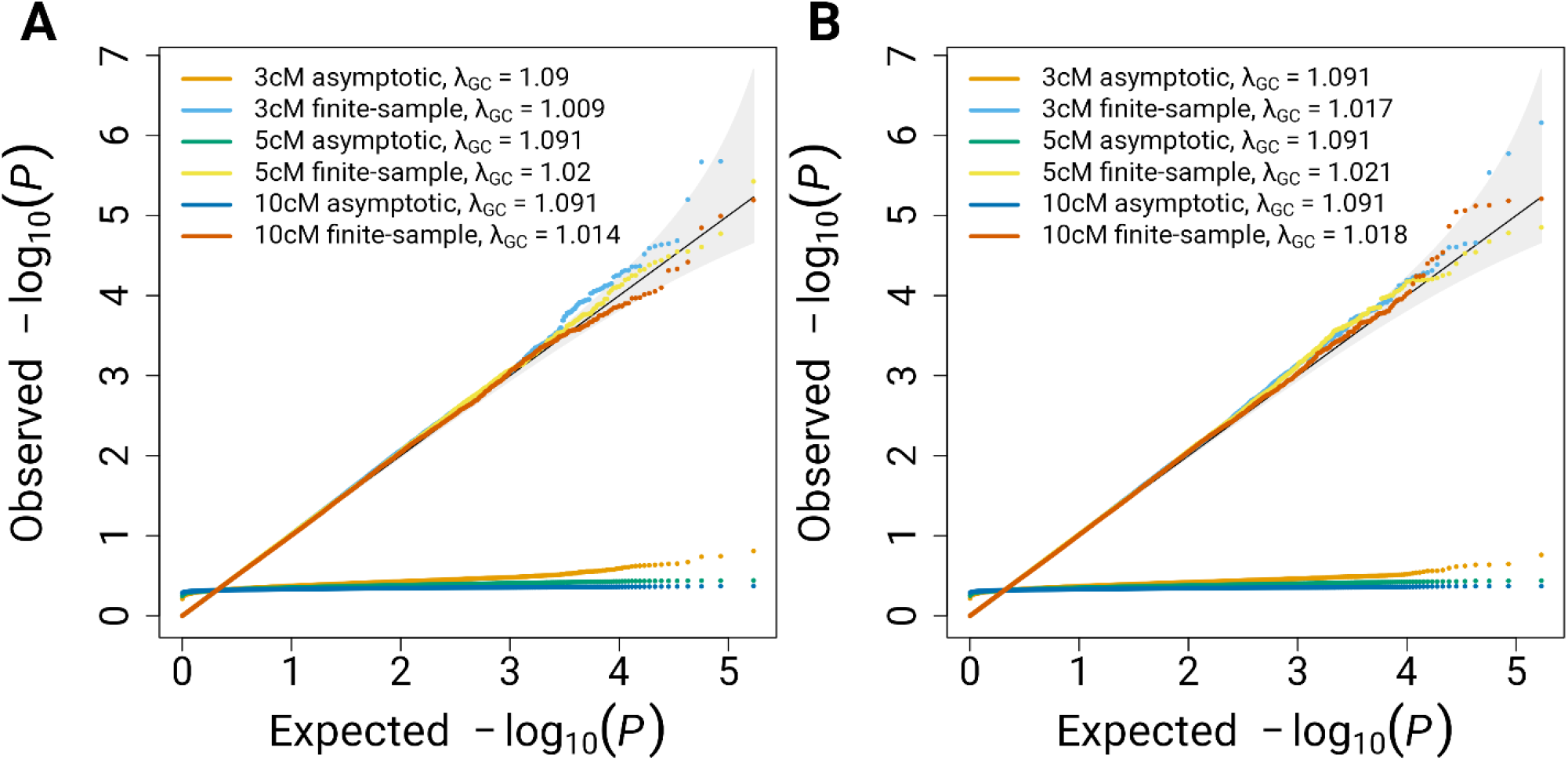
Quantile-quantile plot of asymptotic and finite-sample FiMAP p values under the null hypothesis. A total of 170,150 p values from 50 simulation replicates were plotted. The genomic inflation factor *λ*_*GC*_ was computed using the observed median p values. The local IBD matrices were constructed from (A) RaPID or (B) hap-IBD IBD segments called with length ≥ 3 cM, 5 cM, 10 cM.

### Power

As asymptotic FiMAP p values were extremely conservative in the tail from the type I error simulation studies, we only computed finite-sample FiMAP p values in the power simulation studies as well as the UK Biobank real data analysis. Assuming the causal variants in a 1 cM window were ultra-rare (with MAF < 0.05%) and not directly observed, **Figure 4A** shows that FiMAP with IBD segments called with length ≥ 3 cM and 5 cM outperformed the single-variant test (which was widely used in GWAS) and commonly used variant set tests. Of note, the single-variant test power, even with a Bonferroni correction that multiplied the minimum p value by the number of genotyped variants in the 1 cM window, was higher than the variant set tests with Wu weights^27^ that favored variants with a lower MAF, suggesting that variant set tests may not perform well in the presence of a large number of neutral variants which could dilute the association signal. FiMAP with IBD segments called with length ≥ 10 cM outperformed the burden test but had lower power than the variant set tests SKAT, SKAT-O and SMMAT.

**Figure 4.**
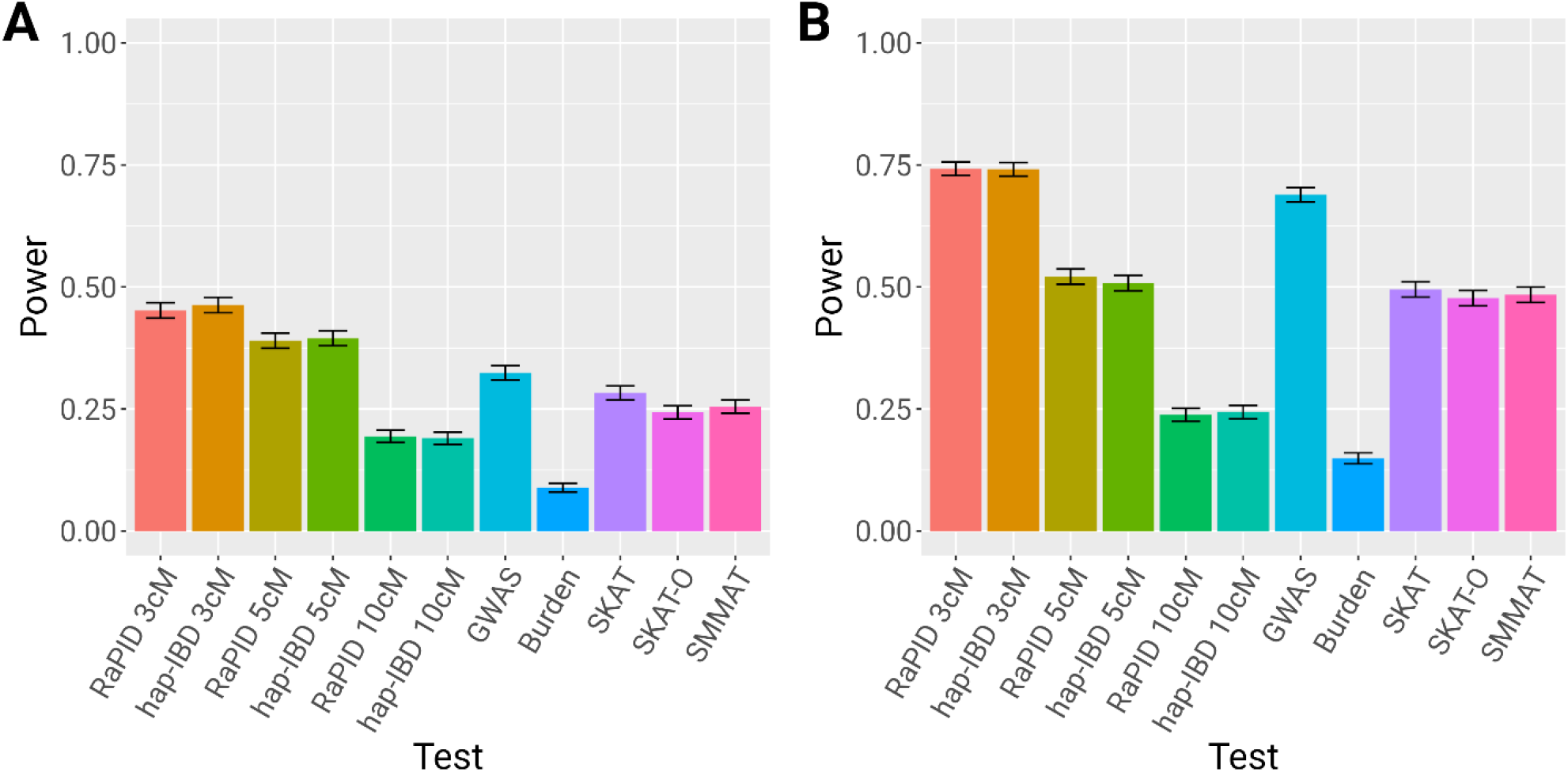
Power comparison of FiMAP, GWAS and variant set tests. (A) Untyped rare causal variants; (B) Causal haplotype effects. In each simulation replicate with 1 cM window, finite-sample FiMAP with local IBD matrices constructed from RaPID and hap-IBD IBD segments called with length ≥ 3 cM, 5 cM, 10 cM were benchmarked with a GWAS single-variant test with a Bonferroni correction for the minimum p value, and four variant set tests on the same window: the burden test, SKAT, SKAT-O and SMMAT. Empirical power was estimated as the proportion of p values less than 0.05 over 1,000 simulation replicates.

In the presence of causal haplotype effects, FiMAP with IBD segments called with length ≥ 3 cM had the best power, followed by the single-variant test and FiMAP with IBD segments called with length ≥ 5 cM (**Figure 4B**). FiMAP with IBD segments called with length ≥ 10 cM suffered from a substantial power loss, likely due to the fact that many shorter IBD segments tagging the causal haplotype in the region were not included. **Figure 4** also shows that in both simulation scenarios, FiMAP power was similar for IBD segments with the same length cutoff, for both RaPID and hap-IBD.

### UK Biobank anthropometric traits

For each anthropometric trait from the UK Biobank, the FiMAP analysis using the RaPID IBD length cutoff 3 cM took 90 CPU hours with a maximum memory footprint scaling linearly with the number of nonzero entries in the local IBD matrix *Ψ*_*l*_. As a comparison, the GWAS of 89 million imputed variants took 1,175 CPU hours on the same computing node for each anthropometric trait. **Table 1** shows that FiMAP using RaPID IBD segments with length ≥ 3 cM identified 1,914 association signals outside of any significant GWAS hits. On the other hand, it also shows that many significant 1 cM windows in FiMAP results had at least one imputed tag variant with MAF ≥ 0.01%, imputation quality score ≥ 0.3, and GWAS p-value < 5×10^−8^ in the same test region. Interestingly, 2,224 out of 3,442 (65%) 1 cM windows remained significant even after a very stringent conditional analysis by adjusting for all independent tag variants with GWAS p-value < 5×10^−8^ in the ± 3 cM flanking regions (a total length of 7 cM), suggesting that IBD mapping provided complementary association evidence missed by a traditional GWAS approach. **Figure 5** and **Figure S1** show that for windows with a significant unconditional FiMAP p-value, conditional p-values were generally larger (less significant) than unconditional p-values as expected, for all 6 anthropometric traits and both RaPID and hap-IBD. Of note, we identified a 6 cM region (29 - 35 cM) on chromosome 21 associated with both standing height and sitting height using RaPID IBD segments with length ≥ 3 cM (**Figure 6**, minimum p values after conditioning on all independent tag variants in each 7 cM region: standing height 9.5 × 10^−19^, sitting height 1.2 × 10^−9^). Alternatively, using hap-IBD IBD segments with length ≥ 3 cM, we identified the same region on chromosome 21 with minimum conditional p value 1.0 × 10^−15^ for standing height and 1.7 × 10^−8^ for sitting height (**Figure S2**).

**Table 1.** Numbers of significant 1 cM windows in UK Biobank unconditional and conditional IBD mapping for 6 anthropometric traits. IBD segments called by RaPID and hap-IBD with length ≥ 3 cM were used in the FiMAP analysis. Statistical significance was defined as p value < 1.47 × 10^−5^ for both unconditional and conditional FiMAP, and single-variant test p value < 5 × 10^−8^ for GWAS. Overlapping with GWAS hits was defined as a significant 1 cM unconditional FiMAP test region with at least one imputed tag variant with MAF ≥ 0.01%, imputation quality score ≥ 0.3, and GWAS p-value < 5×10^−8^ in the same 1 cM test region. Conditional FiMAP tests were performed after adjusting for all independent tag variants with MAF ≥ 0.01%, imputation quality score ≥ 0.3, and GWAS p-value < 5×10^−8^ in the ± 3 cM flanking regions (a total length of 7 cM) from GWAS.

| Trait | Unconditional |  | No GWAS overlaps |  | Conditional |  |
| --- | --- | --- | --- | --- | --- | --- |
|  | RaPID | hap-IBD | RaPID | hap-IBD | RaPID | hap-IBD |
| Waist circumference | 99 | 96 | 66 (67%) | 63 (66%) | 62 (63%) | 49 (51%) |
| Hip circumference | 74 | 65 | 42 (57%) | 36 (55%) | 21 (28%) | 21 (32%) |
| Standing height | 2,189 | 2,266 | 1,242 (57%) | 1,307 (58%) | 1,692 (77%) | 1,773 (78%) |
| Sitting height | 842 | 871 | 443 (53%) | 460 (53%) | 372 (44%) | 418 (48%) |
| Body mass index | 138 | 144 | 78 (57%) | 85 (59%) | 53 (38%) | 62 (43%) |
| Body weight | 100 | 90 | 43 (43%) | 39 (43%) | 24 (24%) | 32 (36%) |

**Figure 5.**
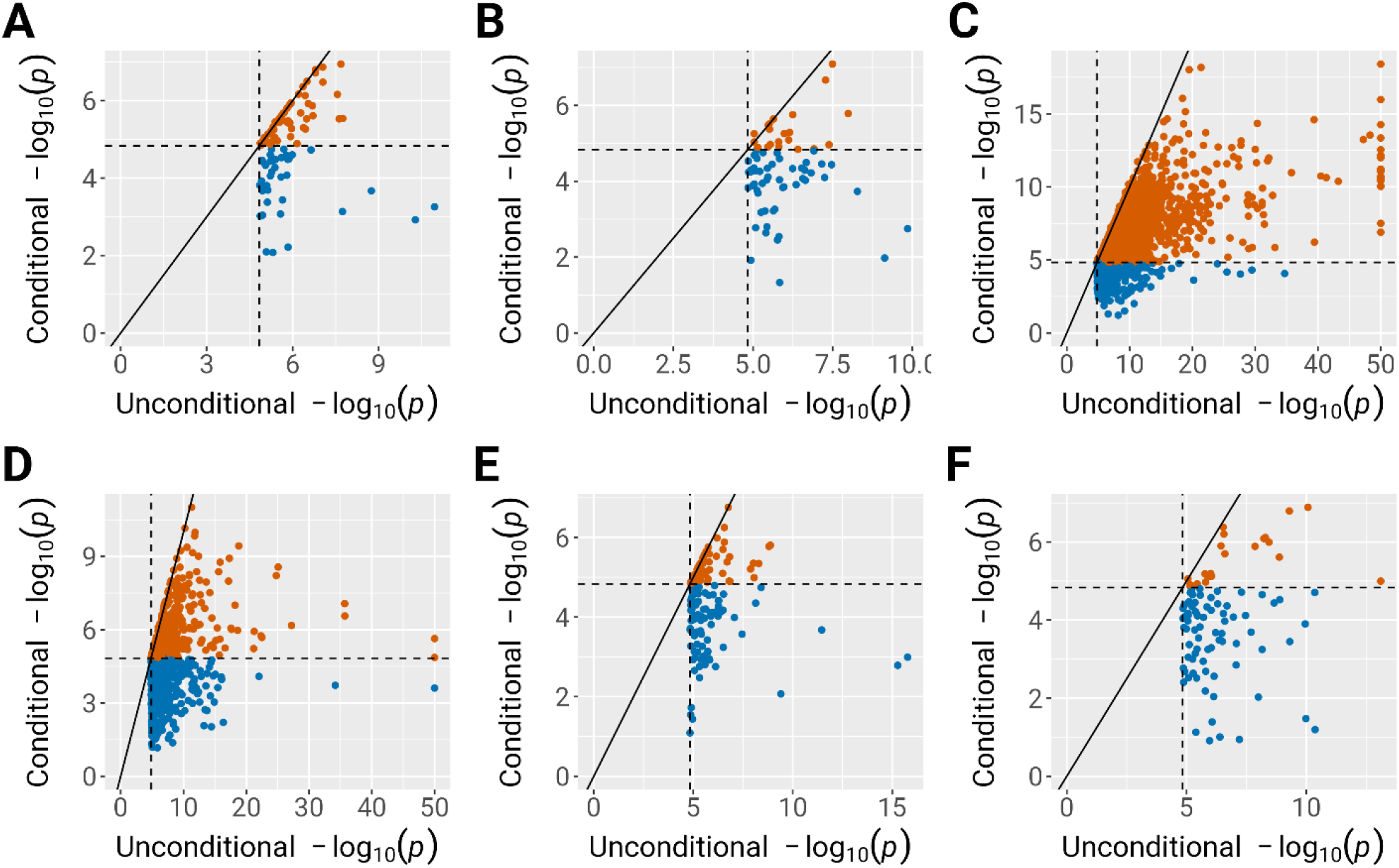
Comparison of FiMAP p values from RaPID IBD segments before and after conditioning on significant tag variants in the testing window and flanking regions. (A) Waist circumference; (B) Hip circumference; (C) Standing height; (D) Sitting height; (E) Body mass index; (F) Body weight. IBD segments called by RaPID with length ≥ 3 cM were used in the FiMAP analysis. Dashed lines represented the Bonferroni-corrected significance level of 0.05/3,403 = 1.47 × 10^−5^. Only significant unconditional FiMAP p values were shown. P values < 1 × 10^−50^ for standing and sitting height were truncated at 1 × 10^−50^. FiMAP conditional p-values that no longer reach significance were shown in blue.

**Figure 6.**
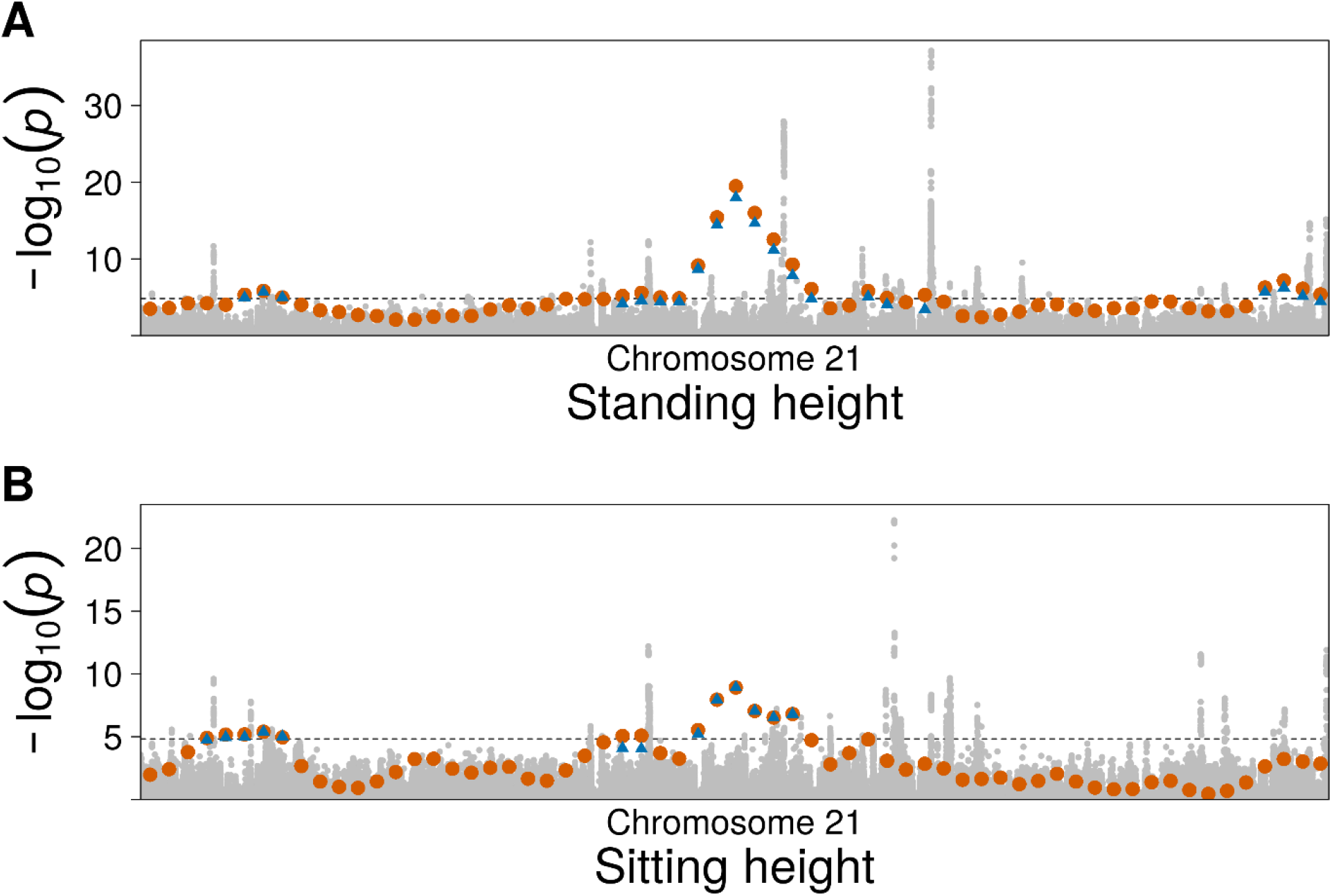
GWAS and FiMAP p values from RaPID IBD segments on chromosome 21. (A) Standing height; (B) Sitting height. IBD segments called by RaPID with length ≥ 3 cM were used in the FiMAP analysis. GWAS p values were shown in grey and unconditional FiMAP p values were shown in orange. For windows with unconditional FiMAP p values < 1.47 × 10^−5^, conditional p values after adjusting for all independent GWAS tag variants were shown in blue triangles.

For four anthropometric traits related to body fat distribution or weight (waist circumference, hip circumference, body mass index, and body weight), we found highly significant FiMAP p values near the *FTO* region on chromosome 16, using both RaPID and hap-IBD IBD segments with length ≥ 3 cM. However, none of the conditional p values remained significant after adjusting for GWAS tag variants in this region, suggesting that FiMAP did not provide additional association evidence beyond GWAS in this case (**Figure S3** and **Figure S4**).

Moreover, on the genome-wide scale, FiMAP p values from RaPID IBD segments with length ≥ 3 cM were highly concordant with those from hap-IBD IBD segments with length ≥ 3 cM (**Figure S5**, minimum Spearman’s *ρ* = 0.913). FiMAP p values from the same IBD caller with different length cutoffs, however, showed much lower correlation (**Figures S6-S9**), suggesting that FiMAP results are much more dependent on the length cutoff than the IBD caller used.

As both RaPID and FiMAP rely on random algorithms, we also used a different random number seed in RaPID IBD segment calling, as well as FiMAP IBD mapping, to investigate the numerical stability of our findings. **Figure S10** and **Figure S11** show that FiMAP p values were highly stable with respect to different random number seeds used in IBD segment calling by RaPID (minimum Spearman’s *ρ* = 0.966) and IBD mapping (minimum Spearman’s *ρ* = 0.989). These results suggested that our top findings were robust against different random number seeds used in IBD segment calling and/or IBD mapping.

## Discussion

In this work, we have developed FiMAP to conduct fast IBD mapping analysis that scales linearly with the sample size and applied it to the UK Biobank data. Compared to traditional GWAS approaches and variant set tests for sequencing data that utilize only unphased genotypes, IBD mapping with the phase information offers a distinct perspective into the genetic architecture of complex traits by leveraging the genotype similarity in a specific genomic region. The heuristics that if a genomic region is associated with a complex trait, then individuals with greater genotype similarity are also expected to show greater phenotype similarity, have led to the development of linkage analysis methods for family data, including the variance component linkage analysis. However, such methods have not previously been widely applied to biobank-scale population-based cohorts, due to computational challenges.

FiMAP is an accurate and computationally efficient IBD mapping method. It leverages a random matrix to approximate the null distribution of the variance component test statistic in quadratic form. We have found in the UK Biobank data analysis that different random number seeds used in either IBD segment calling by RaPID or IBD mapping by FiMAP had minimal impact on the top association findings, as the p values were highly consistent regardless of the different random number seeds. In addition, top association findings from two different IBD callers RaPID and hap-IBD were also highly concordant given the same length cutoff, suggesting that FiMAP was robust with respect to which IBD caller was used.

Our simulation studies showed that FiMAP is often more powerful using IBD segments with shorter length cutoffs. However, there is also a tradeoff between statistical power and computational efficiency. With an IBD length cutoff of 10 cM, FiMAP was very computationally efficient, but also had the lowest power in our simulation settings. With an IBD length cutoff of 3 cM, FiMAP was often more powerful than using cutoffs of 5 cM and 10 cM, but the run time also dramatically increased. With even shorter IBD length cutoffs (e.g., 2 cM), a much larger number of IBD segments are expected to be called (see Equation 14 of Palamara *et al*.),^5^ which may decrease the sparsity of local IBD matrices and lead to a substantial increase in both the run time and memory footprint. In our real data analysis, we chose an IBD length of 3 cM to balance between power and computational efficiency.

In general, the power of IBD mapping depends on the number of IBD segments each sample has in a particular genomic window, which grows quadratically with N. For example, with an IBD length cutoff of 3 cM for RaPID, the median number of IBD segments per individual per tested window was 60.4 in the full sample (N = 487,252), compared to 0.124 in a random subset of N = 1,000 individuals. Therefore, we expect FiMAP to have much greater statistical power in biobank-scale cohorts, although the actual power may also depend on other factors such as the genetic architecture of the trait, as well as the degree of relatedness in the samples. On the other hand, when N is much larger than the size of UK Biobank (in a somewhat distant future), the local IBD matrix may not be sparse enough and new method than FiMAP may be needed for efficient IBD mapping.

Surprisingly, asymptotic p values from FiMAP are not well calibrated even with the sample sizes of the UK Biobank. Although the *N* × *N* local IBD matrices are large in biobank-scale cohorts, they are often sparse with *O*(*N*) non-zero off-diagonal elements. As a result, the asymptotic p values from FiMAP are extremely conservative in the tail. In contrast, our simulation studies under the null hypothesis of no genetic association showed that finite-sample FiMAP p values followed a uniform distribution, after accounting for the variability in estimating the residual variance parameter. Therefore, we recommend the use of finite-sample FiMAP p values for all analyses.

FiMAP, or IBD mapping in general, has several advantages compared to traditional GWAS approaches and variant set tests on unphased genotypes. It may better tag rare causal variants, especially when they are not directly genotyped, which is often the case for commonly used genotyping arrays. Although it is unlikely that rare causal variants are not genotyped using the whole genome sequencing technology, as the IBD calling algorithm such as RaPID tolerates mismatches, FiMAP is more robust to genotyping or sequencing errors since each individual genetic variant would not dramatically change the IBD segment calling. Moreover, the phasing information is usually ignored in traditional GWAS approaches and variant set tests, and our simulation studies showed that FiMAP is more powerful in identifying the genetic association in the case of haplotype effects. Our conditional analysis for UK Biobank anthropometric traits showed that many 1 cM windows identified by FiMAP remained significant after adjusting for all GWAS tag variants in the window and flanking regions, suggesting that these associations could not be fully explained by unphased genotypes, and therefore haplotype effects might play an important role in the genetic architecture of complex traits.

FiMAP also has a few limitations. It tests a few thousand 1 cM windows on the human genome, and therefore requires a much less stringent significance level than traditional GWAS approaches which test millions of genetic variants. However, the major limitation of FiMAP is the low resolution of association signals. Follow-up analysis is often needed to identify causal variants or interpret these association findings. For example, we can shorten the window size from 1 cM to the size of genes, and conduct gene-based IBD mapping tests using local IBD matrices defined on each gene and flanking regions. Also, currently FiMAP can only be applied to quantitative traits in a linear mixed model framework, and future work includes extension to binary traits using logistic mixed models, as well as survival traits using Cox mixed models. Nevertheless, FiMAP empowers computationally efficient IBD mapping for complex traits using variance component linkage analysis models with unprecedented sample sizes from biobank-scale cohorts.

## Data Availability

This research has been conducted using the UK Biobank Resource under Application Number 24247. All data collection was done by the UK Biobank prior to this research. The data used in this study can be requested by applying through the UK Biobank Access Management System.

https://www.ukbiobank.ac.uk/enable-your-research/register

## Supplemental Data

Supplemental Data include 11 figures.

## Declaration of Interests

The authors declare no competing interests.

## Acknowledgments

This work was supported by National Institutes of Health grants R00 HL130593 (to H.C.) and R01 HG010086 (to A.N. and D.Z.). This research has been conducted using the UK Biobank Resource under Application Number 24247.

## Web Resources

The URLs for data presented herein are as follows.

FiMAP, https://github.com/hanchenphd/FiMAP

GMMAT, https://github.com/hanchenphd/GMMAT

RaPID, https://github.com/ZhiGroup/RaPID

## Notes

### Competing Interest Statement

The authors have declared no competing interest.

### Author Declarations

This study was approved by the University of Texas Health Science Center at Houston (UTHealth Houston) IRB (Number HSC-SBMI-17-0366).

### Summary of Updates

We included results from a different IBD caller hap-IBD and updated the UK Biobank analysis results using a length cutoff of 3 cM. We also added 9 supplemental figures.

## References

1. Thompson, E.A. (2013). Identity by descent: variation in meiosis, across genomes, and in populations. Genetics 194, 301–326.

2. Balding, D.J., Nichols, R.A. (1994). DNA profile match probability calculation: how to allow for population stratification, relatedness, database selection and single bands. Forensic Sci. Int. 64, 125–140.

3. Kling, D., Tillmar, A. (2019). Forensic genealogy-A comparison of methods to infer distant relationships based on dense SNP data. Forensic. Sci. Int. Genet. 42, 113–124.

4. Albrechtsen, A., Moltke, I., Nielsen, R. (2010). Natural selection and the distribution of identity-by-descent in the human genome. Genetics 186, 295–308.

5. Palamara, P.F., Lencz, T., Darvasi, A., Pe ’er, I. (2012). Length distributions of identity by descent reveal fine-scale demographic history. Am. J. Hum. Genet. 91, 809–822.

6. Han, L., Abney, M. (2013). Using identity by descent estimation with dense genotype data to detect positive selection. Eur. J. Hum. Genet. 21, 205–211.

7. Ramstetter, M.D., Shenoy, S.A., Dyer, T.D., Lehman, D.M., Curran, J.E., Duggirala, R., Blangero, J., Mezey, J.G., Williams, A.L. (2018). Inferring Identical-by-Descent Sharing of Sample Ancestors Promotes High-Resolution Relative Detection. Am. J. Hum. Genet. 103, 30–44.

8. Astle, W., Balding, D.J. (2009). Population Structure and Cryptic Relatedness in Genetic Association Studies. Statistical Science 24, 451–471.

9. Thornton, T.A. (2015). Statistical methods for genome-wide and sequencing association studies of complex traits in related samples. Curr. Protoc. Hum. Genet. 84, 1.28.1–1.28.9.

10. Wang, B., Sverdlov, S., Thompson, E. (2017). Efficient Estimation of Realized Kinship from Single Nucleotide Polymorphism Genotypes. Genetics 205, 1063–1078.

11. Naseri, A., Shi, J., Lin, X., Zhang, S., Zhi, D. (2021). RAFFI: Accurate and fast familial relationship inference in large scale biobank studies using RaPID. PLoS Genet. 17, e1009315.

12. Goldgar, D.E. (1990). Multipoint analysis of human quantitative genetic variation. Am. J. Hum. Genet. 47, 957–967.

13. Amos, C.I. (1994). Robust variance-components approach for assessing genetic linkage in pedigrees. Am. J. Hum. Genet. 54, 535–543.

14. Xu, S., Atchley, W.R. (1995). A random model approach to interval mapping of quantitative trait loci. Genetics 141, 1189–1197.

15. Almasy, L., Blangero, J. (1998). Multipoint quantitative-trait linkage analysis in general pedigrees. Am. J. Hum. Genet. 62, 1198–1211.

16. Houwen, R.H., Baharloo, S., Blankenship, K., Raeymaekers, P., Juyn, J., Sandkuijl, L.A., Freimer, N.B. (1994). Genome screening by searching for shared segments: mapping a gene for benign recurrent intrahepatic cholestasis. Nat. Genet. 8, 380–386.

17. Gusev, A., Kenny, E.E., Lowe, J.K., Salit, J., Saxena, R., Kathiresan, S., Altshuler, D.M., Friedman, J.M., Breslow, J.L., Pe ’er, I. (2011). DASH: a method for identical-by-descent haplotype mapping uncovers association with recent variation. Am. J. Hum. Genet. 88, 706–717.

18. Browning, S.R., Thompson, E.A. (2012). Detecting rare variant associations by identity-by-descent mapping in case-control studies. Genetics 190, 1521–1531.

19. Vacic, V., Ozelius, L.J., Clark, L.N., Bar-Shira, A., Gana-Weisz, M., Gurevich, T., Gusev, A., Kedmi, M., Kenny, E.E., Liu, X. et al. (2014). Genome-wide mapping of IBD segments in an Ashkenazi PD cohort identifies associated haplotypes. Hum. Mol. Genet. 23, 4693–4702.

20. Hsueh, W.C., Nair, A.K., Kobes, S., Chen, P., Goring, H.H.H., Pollin, T.I., Malhotra, A., Knowler, W.C., Baier, L.J., Hanson, R.L. (2017). Identity-by-Descent Mapping Identifies Major Locus for Serum Triglycerides in Amerindians Largely Explained by an APOC3 Founder Mutation. Circ. Cardiovasc. Genet. 10, 10.1161/CIRCGENETICS.117.001809.

21. Westerlind, H., Imrell, K., Ramanujam, R., Myhr, K.M., Celius, E.G., Harbo, H.F., Oturai, A.B., Hamsten, A., Alfredsson, L., Olsson, T. et al. (2015). Identity-by-descent mapping in a Scandinavian multiple sclerosis cohort. Eur. J. Hum. Genet. 23, 688–692.

22. Henden, L., Twine, N.A., Szul, P., McCann, E.P., Nicholson, G.A., Rowe, D.B., Kiernan, M.C., Bauer, D.C., Blair, I.P., Williams, K.L. (2020). Identity by descent analysis identifies founder events and links SOD1 familial and sporadic ALS cases. NPJ Genom. Med. 5, 32–8. eCollection 2020.

23. Morgenthaler, S., Thilly, W.G. (2007). A strategy to discover genes that carry multi-allelic or mono-allelic risk for common diseases: a cohort allelic sums test (CAST). Mutat. Res. 615, 28–56.

24. Li, B., Leal, S.M. (2008). Methods for detecting associations with rare variants for common diseases: application to analysis of sequence data. Am. J. Hum. Genet. 83, 311–321.

25. Madsen, B.E., Browning, S.R. (2009). A groupwise association test for rare mutations using a weighted sum statistic. PLoS Genet. 5, e1000384.

26. Morris, A.P., Zeggini, E. (2010). An evaluation of statistical approaches to rare variant analysis in genetic association studies. Genet. Epidemiol. 34, 188–193.

27. Wu, M.C., Lee, S., Cai, T., Li, Y., Boehnke, M., Lin, X. (2011). Rare-variant association testing for sequencing data with the sequence kernel association test. Am. J. Hum. Genet. 89, 82–93.

28. Lee, S., Wu, M.C., Lin, X. (2012). Optimal tests for rare variant effects in sequencing association studies. Biostatistics 13, 762–775.

29. Sun, J., Zheng, Y., Hsu, L. (2013). A unified mixed-effects model for rare-variant association in sequencing studies. Genet. Epidemiol. 37, 334–344.

30. Loh, P.R., Palamara, P.F., Price, A.L. (2016). Fast and accurate long-range phasing in a UK Biobank cohort. Nat. Genet. 48, 811–816.

31. Loh, P.R., Danecek, P., Palamara, P.F., Fuchsberger, C., A Reshef, Y., K Finucane, H., Schoenherr, S., Forer, L., McCarthy, S., Abecasis, G.R. et al. (2016). Reference-based phasing using the Haplotype Reference Consortium panel. Nat. Genet. 48, 1443–1448.

32. Delaneau, O., Zagury, J.F., Robinson, M.R., Marchini, J.L., Dermitzakis, E.T. (2019). Accurate, scalable and integrative haplotype estimation. Nat. Commun. 10, 5436–y.

33. Naseri, A., Tang, K., Geng, X., Shi, J., Zhang, J., Shakya, P., Liu, X., Zhang, S., Zhi, D. (2021). Personalized genealogical history of UK individuals inferred from biobank-scale IBD segments. BMC Biol. 19, 32–y.

34. Durbin, R. (2014). Efficient haplotype matching and storage using the positional Burrows-Wheeler transform (PBWT). Bioinformatics 30, 1266–1272.

35. Naseri, A., Liu, X., Tang, K., Zhang, S., Zhi, D. (2019). RaPID: ultra-fast, powerful, and accurate detection of segments identical by descent (IBD) in biobank-scale cohorts. Genome Biol. 20, 143–8.

36. Zhou, Y., Browning, S.R., Browning, B.L. (2020). A Fast and Simple Method for Detecting Identity-by-Descent Segments in Large-Scale Data. Am. J. Hum. Genet. 106, 426–437.

37. Freyman, W.A., McManus, K.F., Shringarpure, S.S., Jewett, E.M., Bryc, K., 23 and Me Research Team, Auton, A. (2021). Fast and Robust Identity-by-Descent Inference with the Templated Positional Burrows-Wheeler Transform. Mol. Biol. Evol. 38, 2131–2151.

38. Nait Saada, J., Kalantzis, G., Shyr, D., Cooper, F., Robinson, M., Gusev, A., Palamara, P.F. (2020). Identity-by-descent detection across 487,409 British samples reveals fine scale population structure and ultra-rare variant associations. Nat. Commun. 11, 6130–x.

39. Shemirani, R., Belbin, G.M., Avery, C.L., Kenny, E.E., Gignoux, C.R., Ambite, J.L. (2019). Rapid detection of identity-by-descent tracts for mega-scale datasets. bioRxiv, 749507.

40. Liu, D., Lin, X., Ghosh, D. (2007). Semiparametric regression of multidimensional genetic pathway data: least-squares kernel machines and linear mixed models. Biometrics 63, 1079–1088.

41. Kwee, L.C., Liu, D., Lin, X., Ghosh, D., Epstein, M.P. (2008). A powerful and flexible multilocus association test for quantitative traits. Am. J. Hum. Genet. 82, 386–397.

42. Chen, H., Wang, C., Conomos, M.P., Stilp, A.M., Li, Z., Sofer, T., Szpiro, A.A., Chen, W., Brehm, J.M., Celedon, J.C. et al. (2016). Control for Population Structure and Relatedness for Binary Traits in Genetic Association Studies via Logistic Mixed Models. Am. J. Hum. Genet. 98, 653–666.

43. Chen, J., Chen, W., Zhao, N., Wu, M.C., Schaid, D.J. (2016). Small Sample Kernel Association Tests for Human Genetic and Microbiome Association Studies. Genet. Epidemiol. 40, 5–19.

44. Zhan, X., Xue, L., Zheng, H., Plantinga, A., Wu, M.C., Schaid, D.J., Zhao, N., Chen, J. (2018). A small-sample kernel association test for correlated data with application to microbiome association studies. Genet. Epidemiol. 42, 772–782.

45. Zhai, J., Knox, K., Twigg, H.L., Zhou, H., Zhou, J.J. (2019). Exact variance component tests for longitudinal microbiome studies. Genet. Epidemiol. 43, 250–262.

46. Chen, H., Huffman, J.E., Brody, J.A., Wang, C., Lee, S., Li, Z., Gogarten, S.M., Sofer, T., Bielak, L.F., Bis, J.C. et al. (2019). Efficient Variant Set Mixed Model Association Tests for Continuous and Binary Traits in Large-Scale Whole-Genome Sequencing Studies. Am. J. Hum. Genet. 104, 260–274.

47. Naseri, A., Zhi, D., Zhang, S. (2020). Discovery of runs-of-homozygosity diplotype clusters and their associations with diseases in UK Biobank. medRxiv.

48. Sofer, T., Zheng, X., Gogarten, S.M., Laurie, C.A., Grinde, K., Shaffer, J.R., Shungin, D., O’Connell, J.R., Durazo-Arvizo, R.A., Raffield, L. et al. (2019). A fully adjusted two-stage procedure for rank-normalization in genetic association studies. Genet. Epidemiol.

49. Kichaev, G., Bhatia, G., Loh, P.R., Gazal, S., Burch, K., Freund, M.K., Schoech, A., Pasaniuc, B., Price, A.L. (2019). Leveraging Polygenic Functional Enrichment to Improve GWAS Power. Am. J. Hum. Genet. 104, 65–75.

50. Zhu, Z., Guo, Y., Shi, H., Liu, C.L., Panganiban, R.A., Chung, W., O’Connor, L.J., Himes, B.E., Gazal, S., Hasegawa, K. et al. (2020). Shared genetic and experimental links between obesity-related traits and asthma subtypes in UK Biobank. J. Allergy Clin. Immunol. 145, 537–549.

51. Hoffmann, T.J., Choquet, H., Yin, J., Banda, Y., Kvale, M.N., Glymour, M., Schaefer, C., Risch, N., Jorgenson, E. (2018). A Large Multiethnic Genome-Wide Association Study of Adult Body Mass Index Identifies Novel Loci. Genetics 210, 499–515.

52. Pulit, S.L., Stoneman, C., Morris, A.P., Wood, A.R., Glastonbury, C.A., Tyrrell, J., Yengo, L., Ferreira, T., Marouli, E., Ji, Y. et al. (2019). Meta-analysis of genome-wide association studies for body fat distribution in 694 649 individuals of European ancestry. Hum. Mol. Genet. 28, 166–174.

53. Akiyama, M., Okada, Y., Kanai, M., Takahashi, A., Momozawa, Y., Ikeda, M., Iwata, N., Ikegawa, S., Hirata, M., Matsuda, K. et al. (2017). Genome-wide association study identifies 112 new loci for body mass index in the Japanese population. Nat. Genet. 49, 1458–1467.

54. Shungin, D., Winkler, T.W., Croteau-Chonka, D.C., Ferreira, T., Locke, A.E., Magi, R., Strawbridge, R.J., Pers, T.H., Fischer, K., Justice, A.E. et al. (2015). New genetic loci link adipose and insulin biology to body fat distribution. Nature 518, 187–196.

55. Uchiyama, K., Totsukawa, G., Puhka, M., Kaneko, Y., Jokitalo, E., Dreveny, I., Beuron, F., Zhang, X., Freemont, P., Kondo, H. (2006). p37 is a p97 adaptor required for Golgi and ER biogenesis in interphase and at the end of mitosis. Dev. Cell. 11, 803–816.

56. Norlin, M., Toll, A., Bjorkhem, I., Wikvall, K. (2000). 24-hydroxycholesterol is a substrate for hepatic cholesterol 7alpha-hydroxylase (CYP7A). J. Lipid Res. 41, 1629–1639.

57. Bodin, K., Andersson, U., Rystedt, E., Ellis, E., Norlin, M., Pikuleva, I., Eggertsen, G., Bjorkhem, I., Diczfalusy, U. (2002). Metabolism of 4 beta -hydroxycholesterol in humans. J. Biol. Chem. 277, 31534–31540.

58. Li, T., Chanda, D., Zhang, Y., Choi, H.S., Chiang, J.Y. (2010). Glucose stimulates cholesterol 7alpha-hydroxylase gene transcription in human hepatocytes. J. Lipid Res. 51, 832–842.

59. Noshiro, M., Okuda, K. (1990). Molecular cloning and sequence analysis of cDNA encoding human cholesterol 7 alpha-hydroxylase. FEBS Lett. 268, 137–140.

60. Shinkyo, R., Xu, L., Tallman, K.A., Cheng, Q., Porter, N.A., Guengerich, F.P. (2011). Conversion of 7-dehydrocholesterol to 7-ketocholesterol is catalyzed by human cytochrome P450 7A1 and occurs by direct oxidation without an epoxide intermediate. J. Biol. Chem. 286, 33021–33028.

