## Supplemental Figures for "FiMAP: A Fast Identity-by-Descent Mapping Test for Biobank-scale Cohorts"

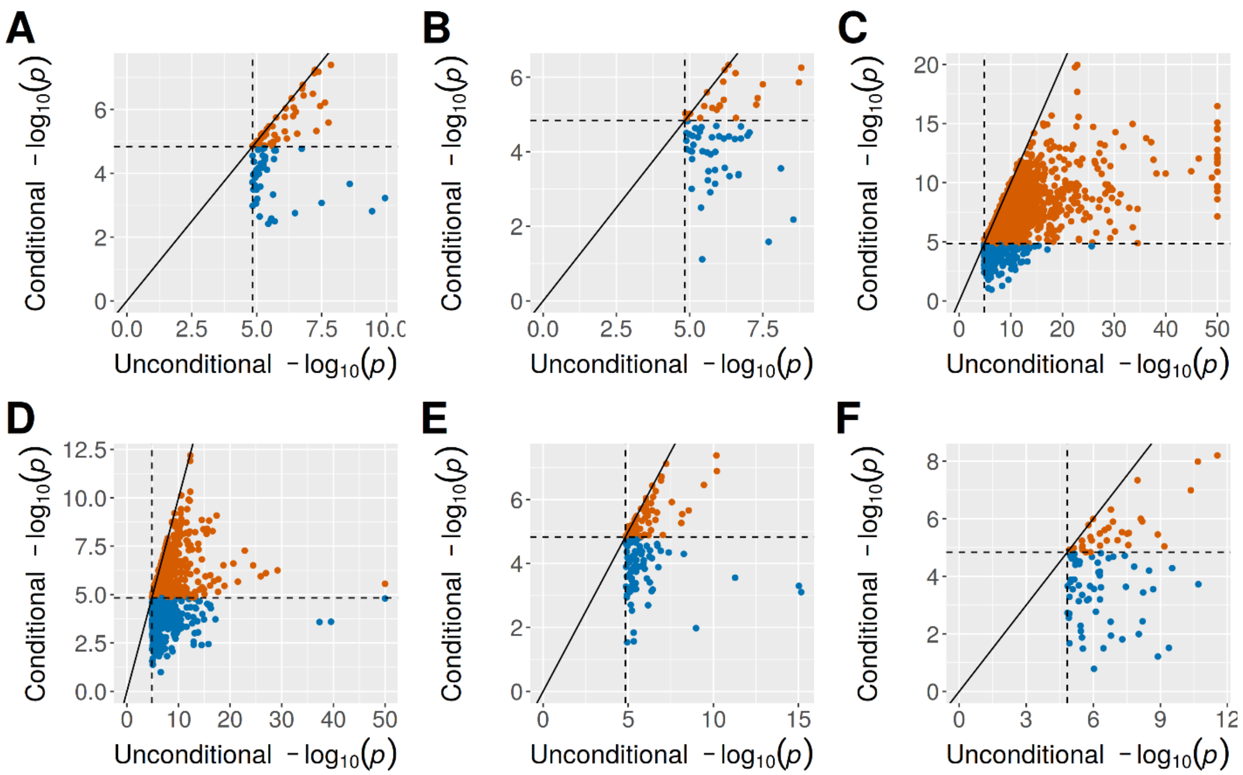


**Figure S1. Comparison of FiMAP p values from hap-IBD IBD segments before and after conditioning on significant tag variants in the testing window and flanking regions.** (A) Waist circumference; (B) Hip circumference; (C) Standing height; (D) Sitting height; (E) Body mass index; (F) Body weight. IBD segments called by hap-IBD with length ≥ 3 cM were used in the FiMAP analysis. Dashed lines represented the Bonferroni-corrected significance level of $0.05/3{,}403 = 1.47 \times 10^{-5}$. Only significant unconditional FiMAP p values were shown. P values < $1 \times 10^{-50}$ for standing and sitting height were truncated at $1 \times 10^{-50}$. FiMAP conditional p-values that no longer reach significance were shown in blue.

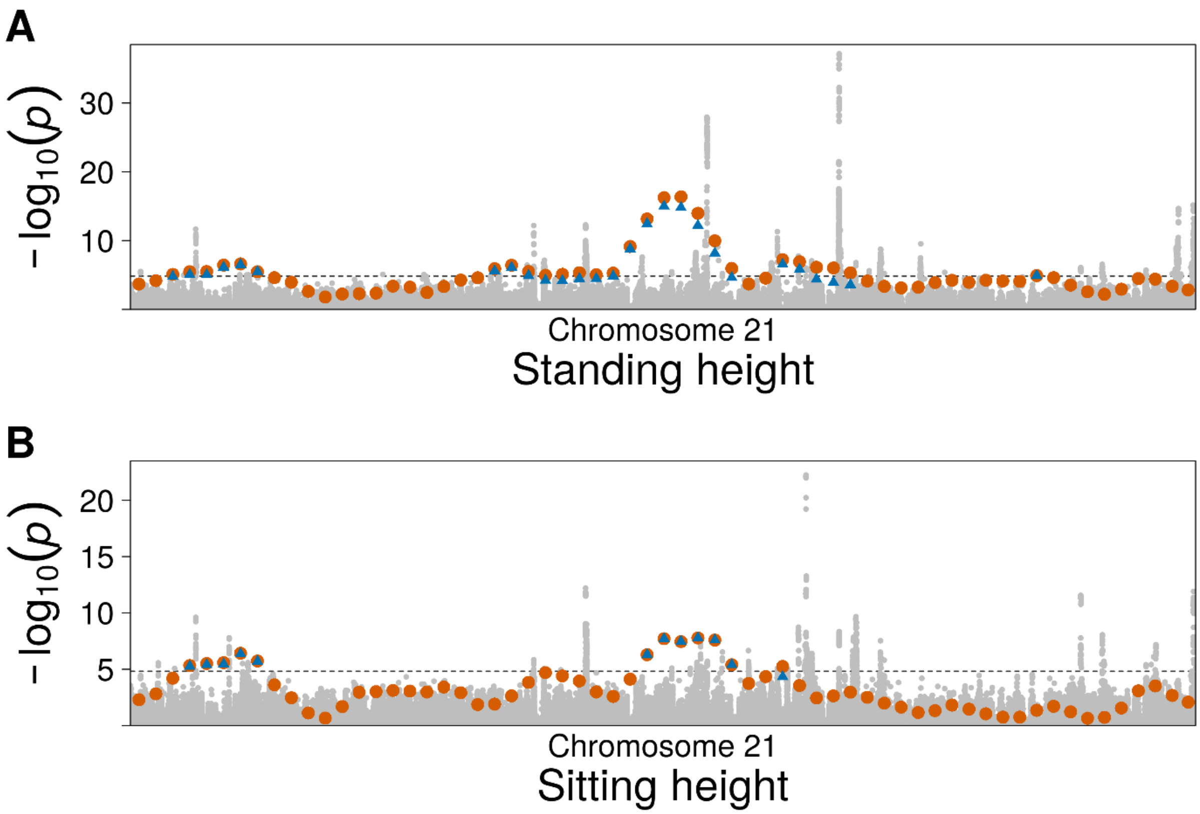


**Figure S2. GWAS and FiMAP p values from hap-IBD IBD segments on chromosome 21.** (A) Standing height; (B) Sitting height. IBD segments called by hap-IBD with length ≥ 3 cM were used in the FiMAP analysis. GWAS p values were shown in grey and unconditional FiMAP p values were shown in orange. For windows with unconditional FiMAP p values $< 1.47 \times 10^{-5}$, conditional p values after adjusting for all independent GWAS tag variants were shown in blue triangles.

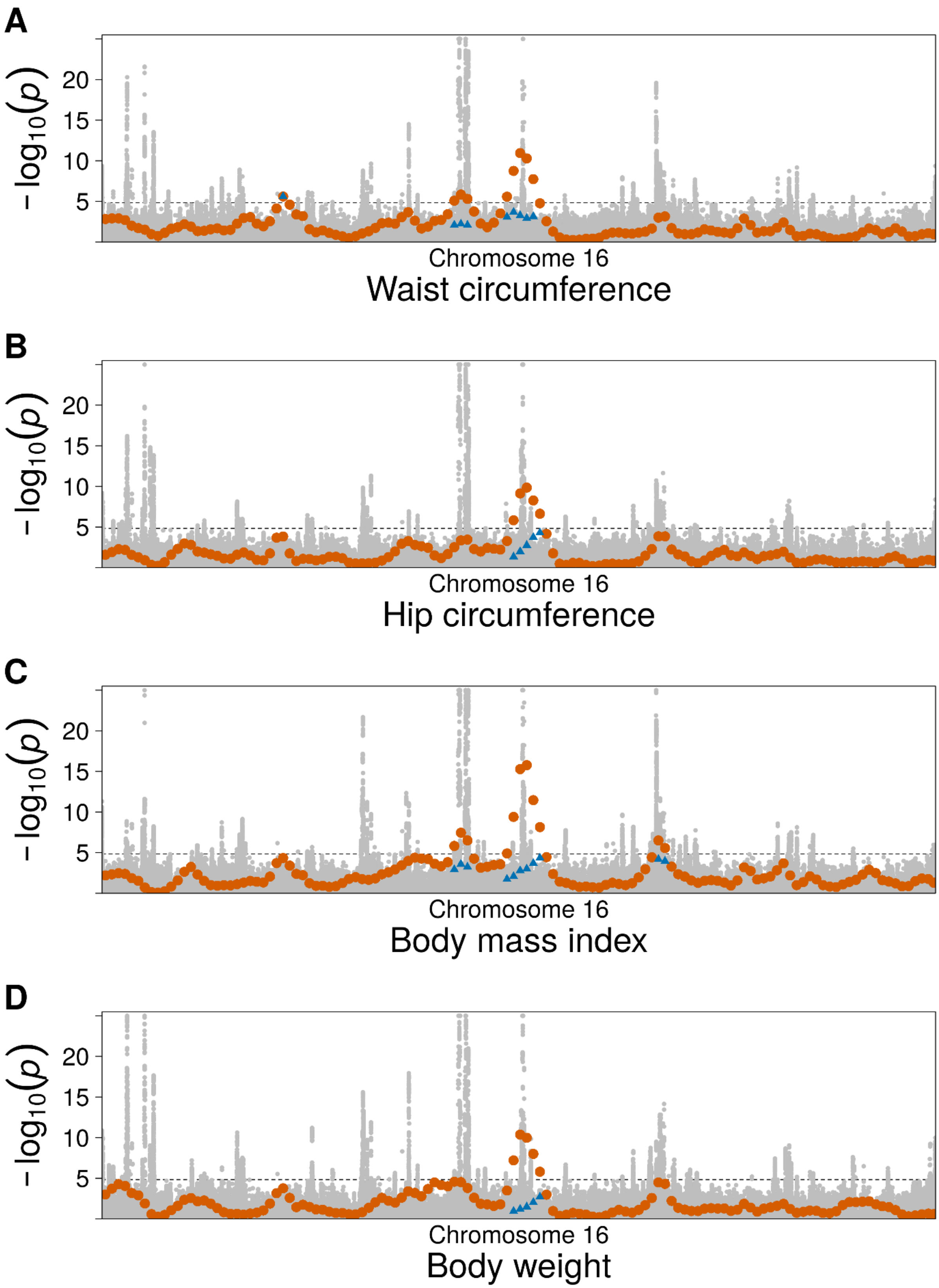
A
B
C
D
−log10(p)
5
10
15
20
Chromosome 16
Waist circumference
Hip circumference
Body mass index
Body weight

**Figure S3. GWAS and FiMAP p values from RaPID IBD segments on chromosome 16.** (A) Waist circumference; (B) Hip circumference; (C) Body mass index; (D) Body weight. IBD segments called by RaPID with length ≥ 3 cM were used in the FiMAP analysis. GWAS p values were shown in grey and unconditional FiMAP p values were shown in orange. GWAS p values $< 1 \times 10^{-25}$ were truncated at $1 \times 10^{-25}$. For windows with unconditional FiMAP p values $< 1.47 \times 10^{-5}$, conditional p values after adjusting for all independent GWAS tag variants were shown in blue triangles.

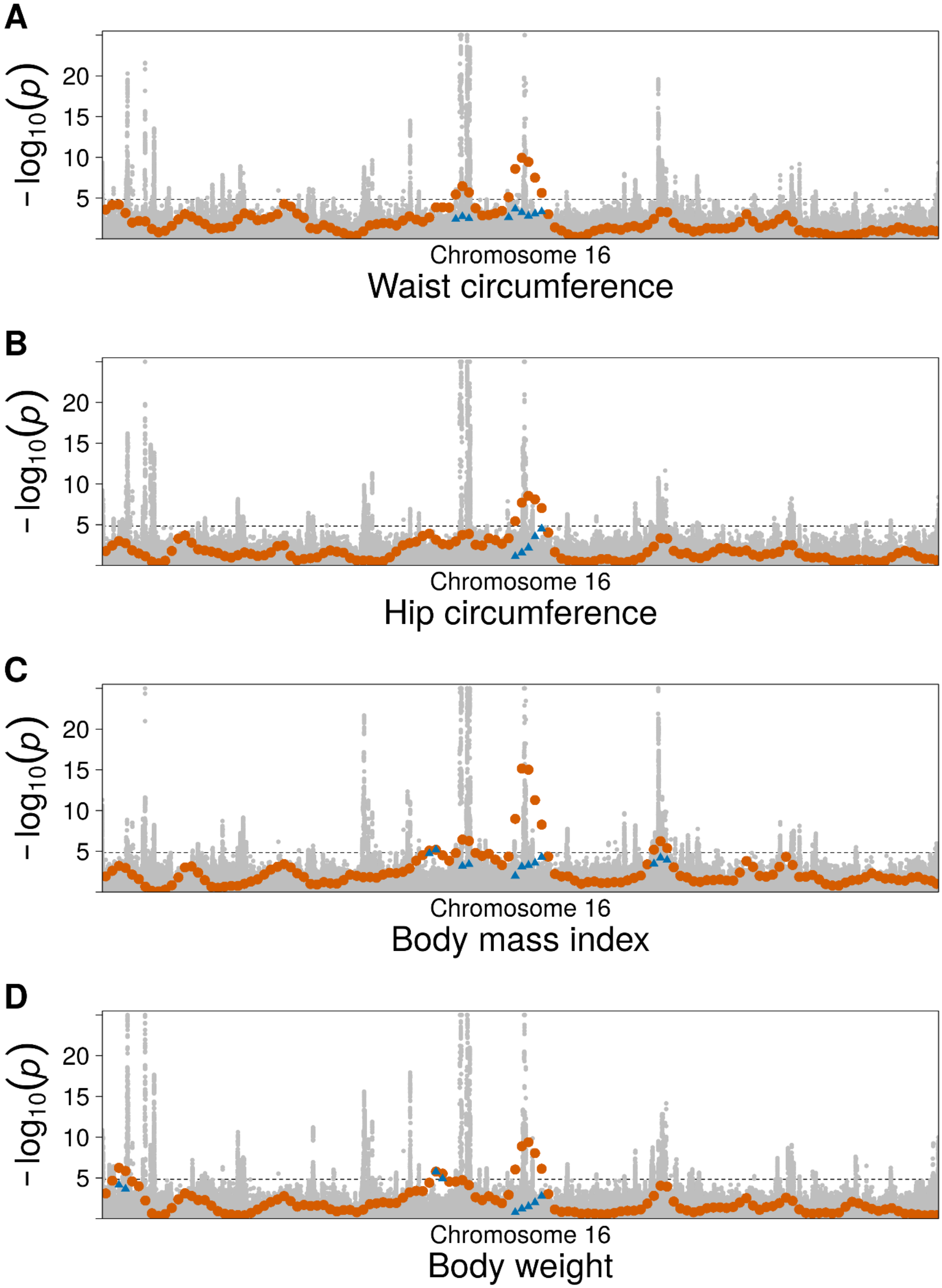
A
$-\log_{10}(p)$
20
15
10
5
Chromosome 16
Waist circumference
B
$-\log_{10}(p)$
20
15
10
5
Chromosome 16
Hip circumference
C
$-\log_{10}(p)$
20
15
10
5
Chromosome 16
Body mass index
D
$-\log_{10}(p)$
20
15
10
5
Chromosome 16
Body weight

**Figure S4. GWAS and FiMAP p values from hap-IBD IBD segments on chromosome 16.** (A) Waist circumference; (B) Hip circumference; (C) Body mass index; (D) Body weight. IBD segments called by hap-IBD with length ≥ 3 cM were used in the FiMAP analysis. GWAS p values were shown in grey and unconditional FiMAP p values were shown in orange. GWAS p values < $1 \times 10^{-25}$ were truncated at $1 \times 10^{-25}$. For windows with unconditional FiMAP p values < $1.47 \times 10^{-5}$, conditional p values after adjusting for all independent GWAS tag variants were shown in blue triangles.

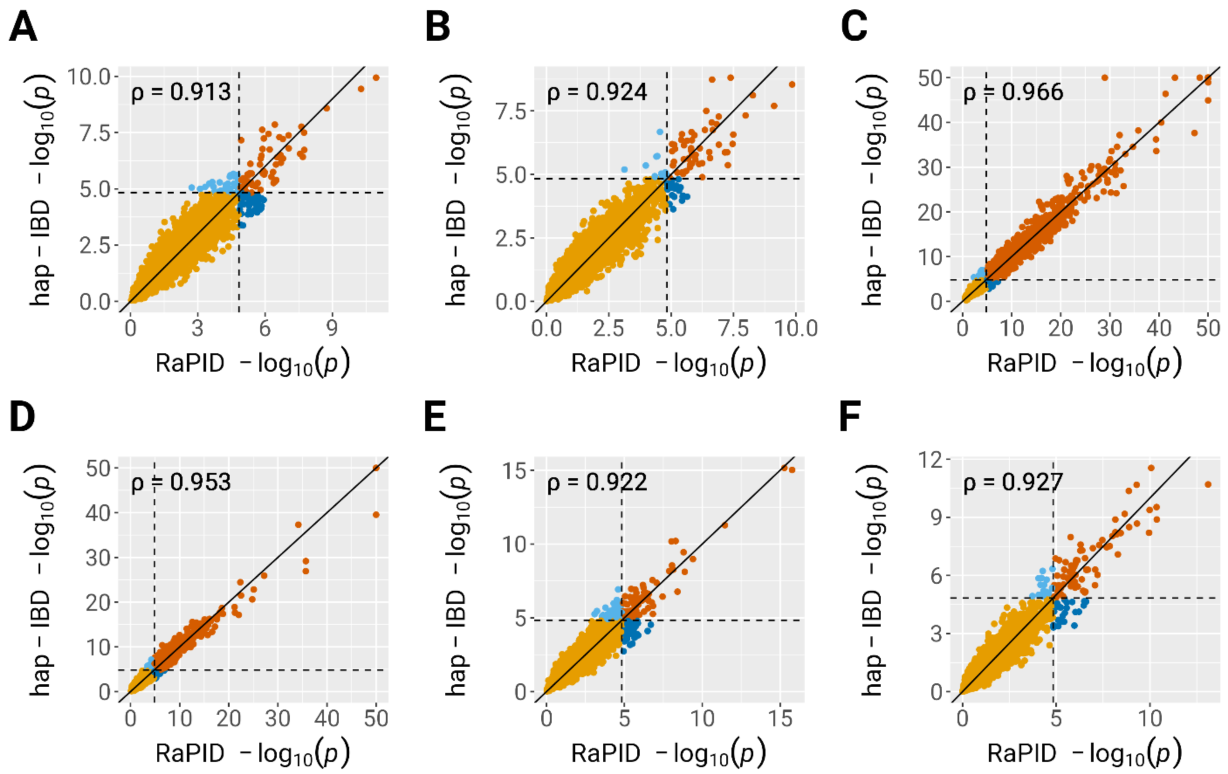


**Figure S5. Comparison of FiMAP p values using RaPID and hap-IBD IBD segments.** (A) Waist circumference; (B) Hip circumference; (C) Standing height; (D) Sitting height; (E) Body mass index; (F) Body weight. IBD segments called by RaPID and hap-IBD with length ≥ 3 cM were used in the FiMAP analysis. Black dashed lines represented the Bonferroni-corrected significance level of 0.05/3,403 = $1.47 \times 10^{-5}$. P values < $1 \times 10^{-50}$ for standing and sitting height were truncated at 1 × 10$^{-50}$. Spearman's rank correlation coefficient was calculated for p values from two separate runs.

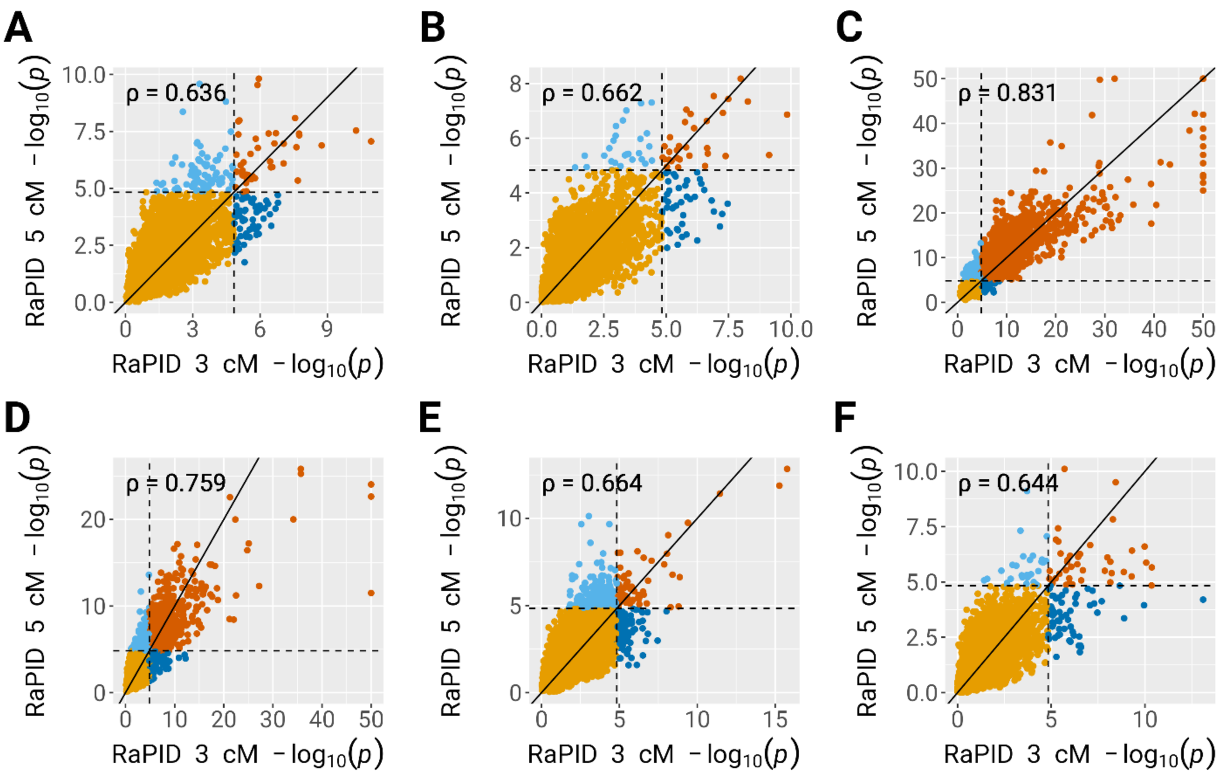


**Figure S6. Comparison of FiMAP p values using RaPID IBD segments with length cutoffs 3 cM and 5 cM.** (A) Waist circumference; (B) Hip circumference; (C) Standing height; (D) Sitting height; (E) Body mass index; (F) Body weight. IBD segments called by RaPID with length ≥ 3 cM and 5 cM were used in the FiMAP analysis. Black dashed lines represented the Bonferroni-corrected significance level of 0.05/3,403 = 1.47 × 10$^{-5}$. P values < 1 × 10$^{-50}$ for standing and sitting height were truncated at 1 × 10$^{-50}$. Spearman's rank correlation coefficient was calculated for p values from two separate runs.

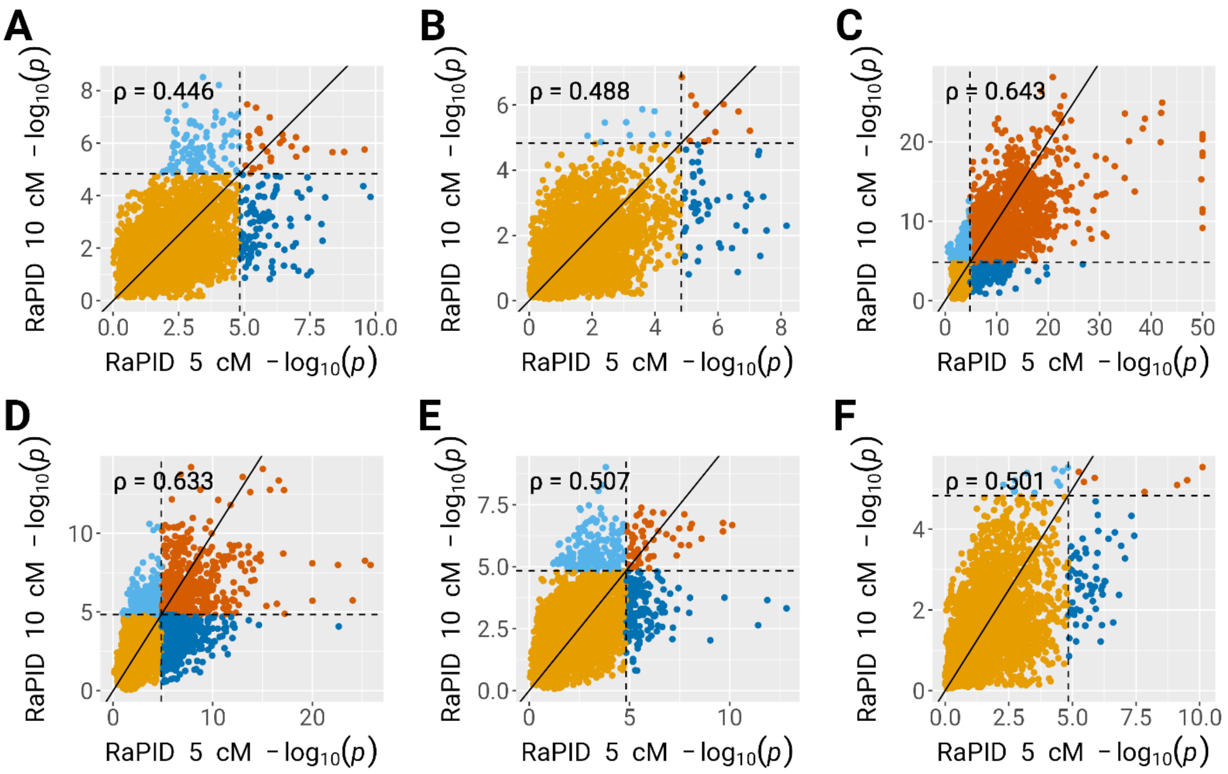


**Figure S7. Comparison of FiMAP p values using RaPID IBD segments with length cutoffs 5 cM and 10 cM.** (A) Waist circumference; (B) Hip circumference; (C) Standing height; (D) Sitting height; (E) Body mass index; (F) Body weight. IBD segments called by RaPID with length ≥ 5 cM and 10 cM were used in the FiMAP analysis. Black dashed lines represented the Bonferroni-corrected significance level of 0.05/3,403 = $1.47 \times 10^{-5}$. P values < $1 \times 10^{-50}$ for standing and sitting height were truncated at $1 \times 10^{-50}$. Spearman's rank correlation coefficient was calculated for p values from two separate runs.

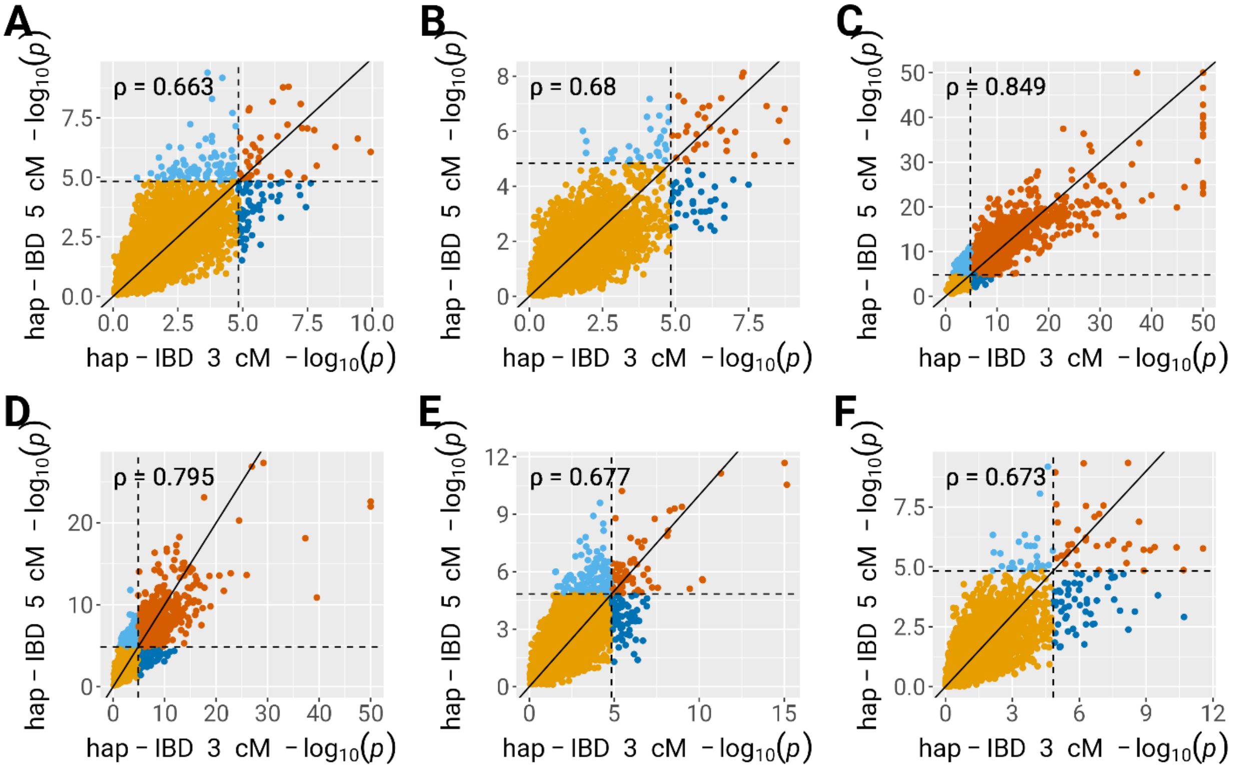


**Figure S8. Comparison of FiMAP p values using hap-IBD IBD segments with length cutoffs 3 cM and 5 cM.** (A) Waist circumference; (B) Hip circumference; (C) Standing height; (D) Sitting height; (E) Body mass index; (F) Body weight. IBD segments called by hap-IBD with length ≥ 3 cM and 5 cM were used in the FiMAP analysis. Black dashed lines represented the Bonferroni-corrected significance level of 0.05/3,403 = $1.47 \times 10^{-5}$. P values < $1 \times 10^{-50}$ for standing and sitting height were truncated at $1 \times 10^{-50}$. Spearman's rank correlation coefficient was calculated for p values from two separate runs.

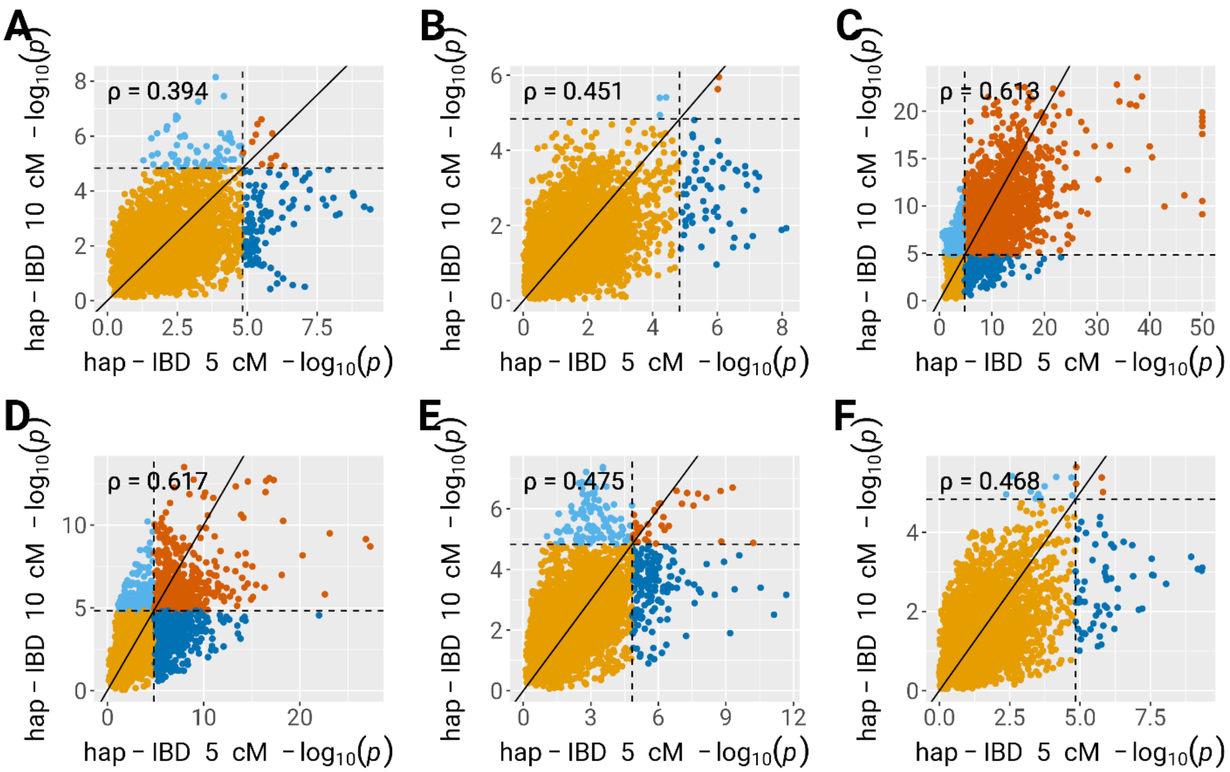


**Figure S9. Comparison of FiMAP p values using hap-IBD IBD segments with length cutoffs 5 cM and 10 cM.** (A) Waist circumference; (B) Hip circumference; (C) Standing height; (D) Sitting height; (E) Body mass index; (F) Body weight. IBD segments called by hap-IBD with length ≥ 5 cM and 10 cM were used in the FiMAP analysis. Black dashed lines represented the Bonferroni-corrected significance level of 0.05/3,403 = $1.47 \times 10^{-5}$. P values < $1 \times 10^{-50}$ for standing and sitting height were truncated at $1 \times 10^{-50}$. Spearman's rank correlation coefficient was calculated for p values from two separate runs.

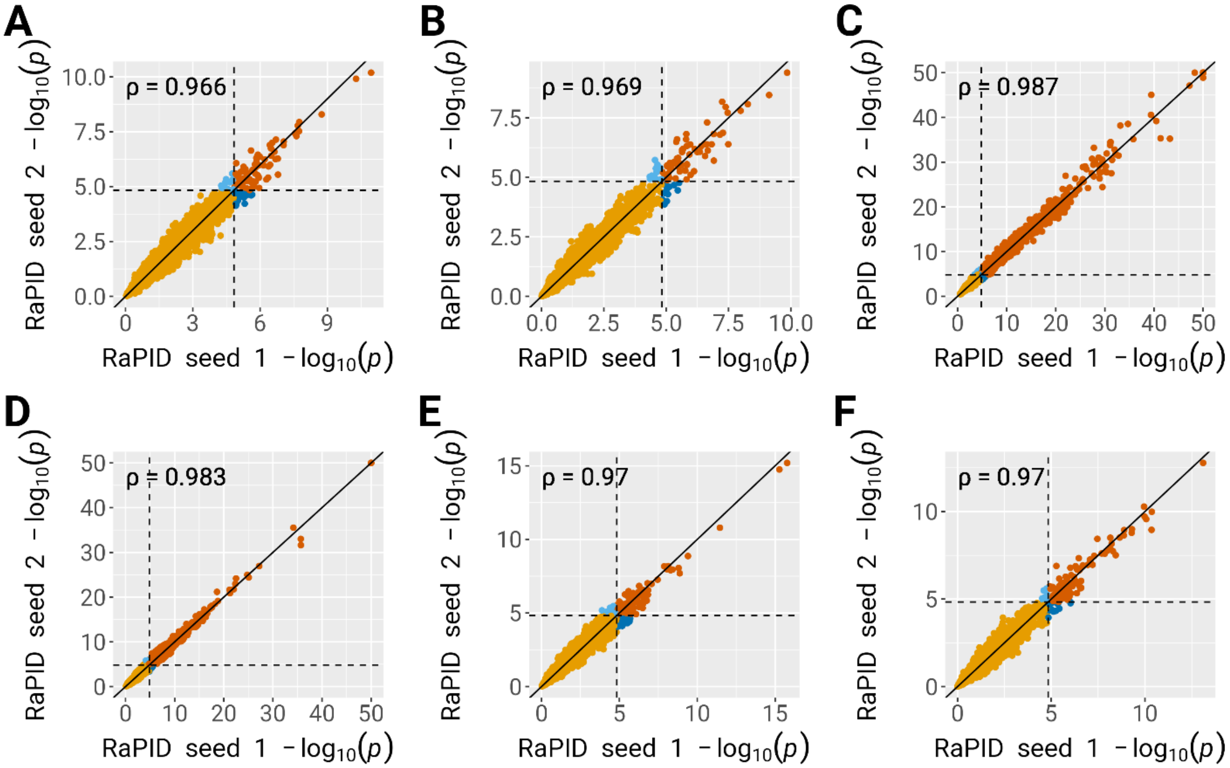


**Figure S10. Comparison of FiMAP p values using two separate RaPID IBD segment calls with different random number seeds.** (A) Waist circumference; (B) Hip circumference; (C) Standing height; (D) Sitting height; (E) Body mass index; (F) Body weight. IBD segments called by RaPID with length ≥ 3 cM were used in the FiMAP analysis. Black dashed lines represented the Bonferroni-corrected significance level of $0.05/3{,}403 = 1.47 \times 10^{-5}$. P values $< 1 \times 10^{-50}$ for standing and sitting height were truncated at $1 \times 10^{-50}$. Spearman's rank correlation coefficient was calculated for p values from two separate runs.

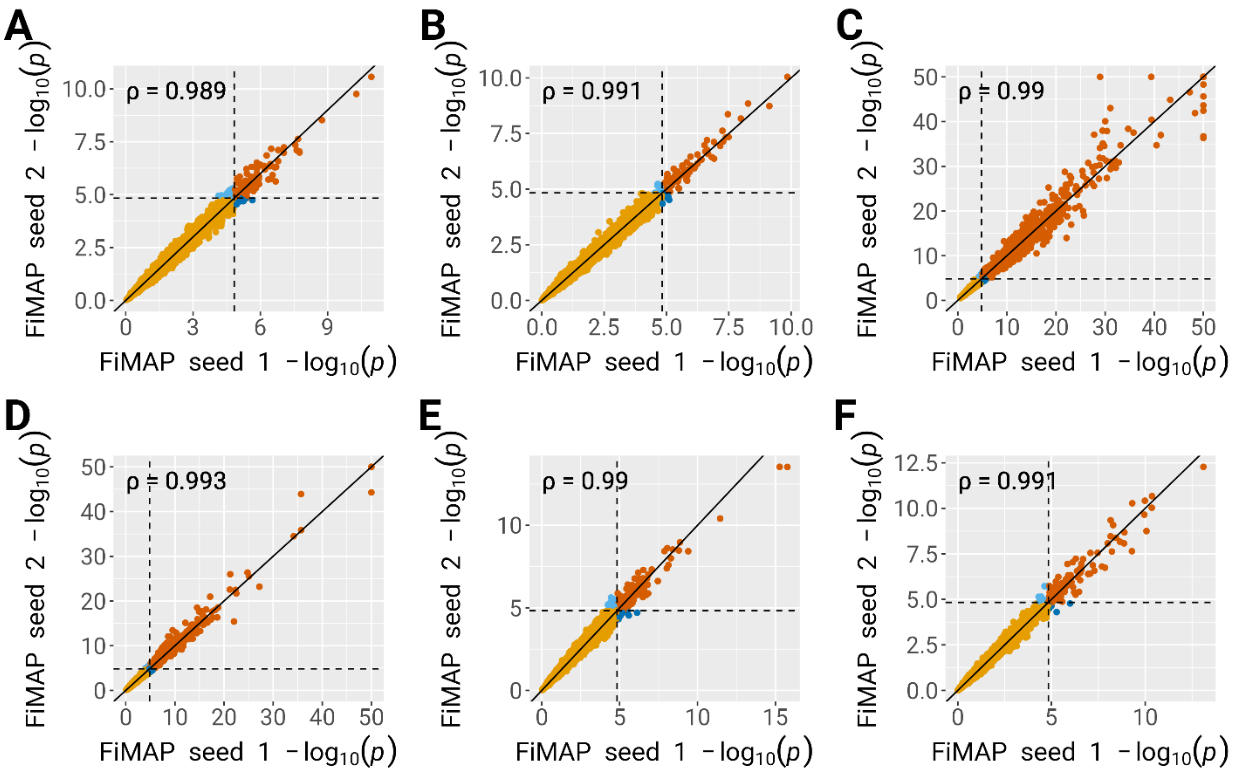


**Figure S11. Comparison of FiMAP p values using two random matrices with different random number seeds.** (A) Waist circumference; (B) Hip circumference; (C) Standing height; (D) Sitting height; (E) Body mass index; (F) Body weight. IBD segments called by RaPID with length ≥ 3 cM were used in the FiMAP analysis. Black dashed lines represented the Bonferroni-corrected significance level of 0.05/3,403 = $1.47 \times 10^{-5}$. P values < $1 \times 10^{-50}$ for standing and sitting height were truncated at $1 \times 10^{-50}$. Spearman's rank correlation coefficient was calculated for p values from two separate runs.
